# On the Role of Financial Support Programs in Mitigating the Sars-CoV-2 Spread in Brazil

**DOI:** 10.1101/2021.11.30.21267063

**Authors:** Vinicius V. L. Albani, Roseane Albani, Nara Bobko, Eduardo Massad, Jorge P. Zubelli

## Abstract

We calculate the impact of a socioeconomic program during 2020 as a measure to mitigate the Coronavirus Disease 2019 (COVID-19) outbreak in Brazil. For each Brazilian State, we estimate the time-dependent reproduction number from daily reports of COVID-19 infections and deaths using a Susceptible-Exposed-Infected-Recovered-like (SEIR-like) model. Then, we analyse the correlations between the reproduction number, the amount of individuals receiving governmental aid, and the index of social isolation based on mobile phone information. We conclude that socioeconomic programs had a significant impact on reducing the accumulated numbers of infections and deaths by allowing those in need to stay at home, adhering to social isolation.

## 1 Introduction

Coronavirus Disease 2019 (COVID-19) pandemic has challenged public authorities on a series of perspectives including public health and the economy. By 10-Nov-2021, COVID-19 caused more than 250 million infections and 5.1 million deaths [1]. During 2020, as vaccination campaigns and vaccination roll-out plans only started by the end of the year in some countries, the only possible disease contention measures, besides social distance and the use of face masks, were lockdowns. Lockdowns were widely used and numerous countries have succeeded to control and reduce COVID-19 infections. However, the use of such kind of measure impacted the economy and, in 2020, the International Monetary Fund have estimated a drop of 3.5% in the world output [2]. In order to reduce economic losses and save the economic activity, many countries have implemented a series of programs to fund individuals that lost their jobs during lockdowns and to help endangered businesses to survive.

In the whole text we converted the amounts in BRL to USD, using the conversion rate of $1,000.00 BRL = $182.72 USD from 10-Nov-2021 [3].

In Brazil, a massive social program called “*Auxílio Emergencial*” (AE) was created to pay a monthly amount to eligible individuals from $54.82 to $219.26 USD. The monthly amount depended on a series of factors, as the number of family members and the period of the year. In 2020, the program was operational from April to December, then it was suspended in January to March 2021, returning in April 2021, and, by June 2021, it was still in function but paying lower monthly amounts, ranging from $27.41 USD to $68.52 USD, with mean value of $45.68 USD [4]. The program reached a median proportion of 22.2% (min–max 14.7%–24.1%) of the Brazilian population, paying the monthly mean amount of $154.86 and $101.81 USD during April to August and September to December 2020, respectively [5]. The monthly payments represented a substantial proportion of the Brazilian minimum wage in 2020, which was $190.94 USD [6], and the total amount invested in the program represented 3.88% of the Brazilian gross domestic product (GDP) in 2020 [7].

During the COVID-19 pandemic, different studies investigated the relation of ethnic and socioeconomic characteristics with the risk of developing SARS-CoV-2 infection. For example, Niedzwiedz *et al*. [8] used Poisson regression to study such a relation in the United Kingdom and found that some ethnic minorities and those under socioeconomic deprivation were associated with a higher risk of infection. Similarly, Hoebel *et al*. [9] compared differences in COVID-19 incidence within groups under different levels of socioeconomic deprivation during the second wave in Germany, finding that during the beginning of the wave, the incidence was higher in less deprived locations, but the situation reversed from the middle to the end of the outbreak. They also found that women in locations with high socioeconomic deprivation had a higher risk of infection. In Cape Town, South Africa, Shaw *et al*. [10] investigated the relationship between antibody positivity, COVID-19 symptom status, medical history, and sociodemographic variables using a seroprevalence study dataset. The study concluded that seropositivity was significantly associated with socioeconomic deprivation. Horta *et al*. [11] achieved similar findings, which analyzed COVID-19 incidence in Brazil through a nationwide seroprevalence study undertaken in 2020. The prevalence of antibodies against SARS-CoV-2, according to this study, was highly dependent on socioeconomic status, education, and ethnic group, with the poorest individuals, those with little education, those self-denominated black or brown, as well as indigenous people were at higher risk of infection. Allan-Blitz *et al*. [12], based on results of SARS-CoV-2 tests from testing sites in Los Angeles, United States, and accounting for United States Census report data on average income, healthcare coverage, and employment status by zip code, the authors found that individuals from places with lower average income, lower rates of employment, or lower rates of health insurance were more likely to test positive for COVID-19 infection. Therefore, socioeconomic deprivation seems to be a relevant factor in the risk of developing SARS-CoV-2, which indicates the supporting role of programs like the *Auxílio Emergencial* (or AE) in disease spread control during outbreaks.

In 2020, non-pharmaceutical measures, like lockdowns, were widely used to control COVID-19 outbreaks. Thus, many recent works have investigated the relationship between mobility and disease spread control, using official reports of COVID-19 infections, mobility data from different sources, and several data analysis tools. For example, using statistical methods, Kissler *et al*. [13] compared Facebook mobility data from New York City, United States (US), with SARS-CoV-2 prevalence, finding that the virus prevalence was lower in locations where individuals traveled less to neighbor places. In Larrosa *et al*. [14], official reports of infections and mobility data from Google in Argentina were used to access the effectiveness of lockdowns and social distancing. The authors concluded that such restriction measures were more effective for short-term dissemination spread than in the long-term. Using an agent-based spatial model and mobility data from Facebook in different countries, Kishore *et al*. [15] provided that mobility restrictions effectively reduced the contact between individuals and controlled outbreaks. In Erim *et al*. [16], interrupted time series were applied to analyze mobility data from mobile phones in Nigeria and their association with reports of COVID-19 infections, finding that mobility restrictions were associated with the reduction in the number of cases.

In Rüdiger *et al*. [17], a statistically-based metric of the social-distancing behavior, the so-called *contact index*, was developed using cell phone GPS data from Germany. The authors found a high correlation between the disease incidence and the contact index. Mehta *et al*. [18] Used logistic regression and Monte Carlo Markov Chain Methods to assess the association between holiday meetings and SARS-COV-2 positivity in the weeks following the 2020 Thanksgiving in the US. They concluded that individuals who had guests or traveled more were more likely to test positive if they also had, for example, participated in more than one non-essential activity by day in the prior weeks of the holiday. Candido *et al*. [19] applied a mobility-driven transmission model and found that non-pharmaceutical interventions, such as lockdowns, considerably reduced the SARS-CoV-2 incidence in São Paulo and Rio de Janeiro, Brazil. Bisanzio *et al*. [20] presented a methodology to predict the spatiotemporal dissemination of reported COVID-19 cases at a global level using geolocated Twitter data. Tomori *et al*. [21] quantified the change in social contact patterns and aggregated mobility information in Germany. They concluded that contact survey data seemed to reflect infection dynamics better than population mobility data, and these data can be related to different aspects of infection dynamics.

In Li *et al*. [22], based on community mobility metrics from Google, the authors found that increasing visits to retail and recreation places, workplaces, and transit stations are linked to the increase of the transmission of Sars-Cov-2 in different cities across the UK. Alleman *et al*. [23] proposed an SEIR-like metapopulation model to describe SARS-CoV-2 spread accounting for the disease characteristics and social contact, where social contact patterns are based on Google Community Mobility data and hospitalization data from Belgium. The authors achieved that reopening schools during lockdown could lead to a substantial increase in virus transmission. Lison *et al*. [24] investigated the relation between mobility, based on mobile phone information, and the propagation of the Sars-CoV-2 in Switzerland during the first and second waves. Using a regression model, the authors found that mobility reduction translates into a reduction of the disease spread. In Brown *et al*. [25], mobility measures based on mobile phone data from Canada were used to quantify the mobility level needed to control SARS-CoV-2 spread. The authors found that each 10% increase in the difference between the observed and the mobility level necessary to control propagation was associated with a 25% increase in the weak growth rate of infections. Tobias [26] used quasi-Poisson regression to analyze trends of incident cases, deaths, and intensive care unit admissions in Italy and Spain before and after national lockdowns, concluding that after lockdowns, incidence trends were considerably reduced in both countries. Coelho *et al*. [27] used mobile phone data, before and during the pandemic, to assess the movement patterns between cities within the states of São Paulo and Rio de Janeiro in Brazil. Such patterns were used in the simulations of a spatial-temporal model to predict potential foci of infections. Peixoto *et al*. [28] identified places in Brazil with a higher risk of outbreaks and higher social vulnerability using air travel statistics, demographic information, socioeconomic indicators, health care capacity, and reports of infections. The data was analyzed using probabilistic models and multivariate cluster analysis.

In summary, these articles illustrate that there are strong evidences that mobility restrictions implies in reduction of contacts and in the disease incidence.

This study aims to investigate if a social program, namely the AE, which reached a considerable fraction of the Brazilian population during the COVID-19 pandemic, is an effective tool in disease contention, since other more effective measures, like vaccination, were unavailable in 2020.

The analysis is based on a comparison between different time series for each Brazilian state, that include the so-called social isolation index (SII), which is based on mobile phone data [29], the daily number of reported COVID-19 infections, the time-dependent reproduction number ℛ(*t*), the statewide population proportion receiving the AE, and the monthly mean amount paid by the program. The results are aggregated by the Brazilian regions due to their large geographic, economic, and demographic differences.

Analyzing socioeconomic measures under the public health perspective and estimating its quantitative impact as an outbreak contention policy, to the best of the authors knowledge, are the main contributions of this work. Such methodology sheds light on the important role such polices can play in the future, helping public authorities to design coordinate responses to control epidemics of emerging infectious diseases.

The article is divided as follows, Section 2 presents the epidemiological model and the data analysis methodology. Results are presented and analyzed in Section 3. Section 4 discusses the proposed methodology and concluding remarks are designed in Section 5.

## 2 Methods

Since we aim to analyze and compare the evolution of COVID-19 infections with social isolation index and the AE data, we propose a susceptible-exposed-infected-recovered-like (SEIR-like) model to estimate the disease dynamics from the data of infections and deaths [30], as well as the reproduction number, which is evaluated using the next generation matrix technique [31].

### 2.1 The Epidemiological Model

The SEIR-like model accounts for five compartments, namely, susceptible (S), exposed (E), infective (I), recovered (R), and deceased (D). The dynamics between compartments is given by the following system of ordinary differential equations without demography:

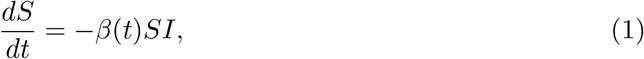

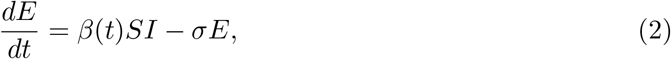

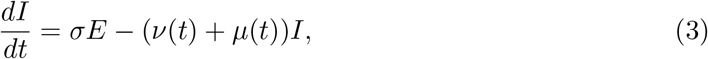

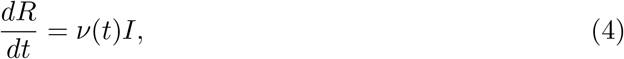

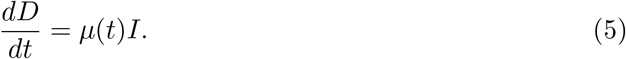

Here, *β*(*t*) denotes the unknown time-dependent infection rate, *σ* represents the inverse of the infection to onset mean time estimated as 5.1 days [32], *ν* is the recovery rate, that we set to 1 *− μ*, where *μ* is the death rate. For each date *t*, we set as the reported deaths on day *t* divided by the reported infections on day *t −* 12, where 12 is the estimated meantime from infection to death [30, 33, 34].

The infection rate for each date *t* is estimated from the daily reported infections by minimizing the following function

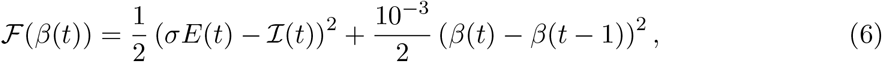

where ℐ (*t*) represents the COVID-19 reports on day *t*.

The objective function in Eq. (6) is minimized to calibrate the transmission parameters of the model in the system in Eqs. (2)–(5). The first part of the right-hand side (RHS) of Eq. (6) evaluates the square of the Euclidean distance between the daily reports of COVID-19 infections, denoted by ℐ (*t*), and their corresponding model predictions, represented by *σE*(*t*). The second part of the RHS of Eq. (6) is the penalty term, that stabilizes the minimization and avoids overfitting. It states that the square of the Euclidean distance between the calibrated *β*(*t*) and *β*(*t−*1) must be minimal. However, the importance of this term in the minimization is stated by the regularization parameter, namely, 10^*−*3^*/*2, which balances the introduction of prior information and the reduction of overfitting [35].

### 2.2 Data Analysis

After calibrating the model and evaluating the reproduction number *ℛ*(*t*), we compare, for each Brazilian State, the following datasets:

1. The daily reported COVID-19 infections during the period 01-Mar-2020 to 11-May-2021 [36].
2. The reproduction number *ℛ*(*t*) obtained from the SEIR-like model for the same period, and its monthly median values.
3. The statewide population proportion receiving the AE from April to December 2020.
4. The mean amount paid monthly by the AE from April to December 2020.
5. The daily social isolation index for the period 01-Mar-2020 to 19-Mar-2021, and its monthly median values.
6. The empirical correlation between the daily changes in the social isolation index and the reproduction number.

We also take into consideration the estimated statewide monthly average income and unemployment in 2020 [37, 38]. We do not consider education indexes since, by 2019, there were no significant variations between them in different Brazilian regions [39]. The analysis is summarized by each Brazilian region.

The correlation evaluation is performed as follows. Firstly, we replaced the original time series of the reproduction number ℛ(*t*) and the social isolation index SII(*t*) by their 7-day moving averaged versions. Then, we evaluated their corresponding daily changes, i.e.,

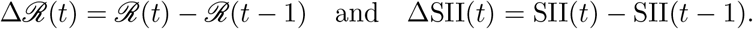

A moving window of 30 consecutive days was used to evaluate the time series of the correlation between Δℛ and ΔSII. We also tested different delays between the series of Δℛ and ΔSII, i.e, the correlations between Δℛ(*t* + Δ*t*) and ΔSII(*t*), with Δ*t* ranging from 0 to 30 days, were evaluated. For each state, the delay in time that led to the maximum number of negative values in the correlation time series was selected. We expect a negative correlation since an increase in the social isolation index, in principle, must cause a drop in the reproduction number. Therefore, this procedure can be interpreted as the mean time that an increment in SII takes to cause a change in ℛ.

### 2.3 Estimating the Impact of the AE on the Number of Infections

To estimate the quantitative impact of the AE on the accumulated numbers of infections, we perform the following steps:

1. Firstly, we consider the estimated delay between changes in the social isolation index and the corresponding changes in the reproduction number.
2. Then, we use such delay to define the estimated transmission parameter *β* as a function of SII. We perform this for all realizations of *β* estimated using bootstrap techniques [40].
3. For each Brazilian state, we aggregate the resulting set of *β* values as functions of SII. This procedure generates a set of possible values for *β* associated with each SII value.
4. We evaluate the number of infections by using the *β* values obtained by changing SII, the SEIR-type model in Eqs. (2)–(5).
5. We exclude the values smaller than the observed ones, evaluate median values, and select the confidence intervals.

## 3 Results

Brazilian states are grouped into five geographic regions, namely, North (N), Northeast (NE), Central-West (CW), Southeast (SE), and South (S), where each of them has intrinsic geographic, economic, and demographic characteristics important for the analysis that follows. For example, the monthly average income varies considerably between these regions, and the impact of the AE on social isolation adherence may differ accordingly. Thus, we analyze the datasets for each state, then we summarize it for each Brazilian region.

Figure 1 and Table 1 present, for each Brazilian State, the accumulated numbers of COVID-19 infections per 100K individuals in 2020, the statewide median population proportions receiving the AE in 2020, the median values of the social isolation index in 2020 and 2021, the statewide monthly average income in 2020, and the statewide unemployment in 2020. Some states, where larger proportions of the population receiving the social-program support, presented larger median values of the social isolation index. In general, States with the higher unemployment and lower monthly average incomes in 2020 presented larger population proportions covered by the AE. Moreover, besides exceptions, states with larger accumulated numbers of COVID-19 per 100K individuals were the same that presented lower SII values. The summary for each region can be found below. Additional figures and tables are in the Supplementary Material.

**Table 1:** Total COVID-19 infections and deaths per 100K individuals reported in 2020 (Cases and Deaths), median, minimum and maximum monthly mean values paid by AE in 2020 (Auxílio 2020), median and 75% confidence interval of the daily social isolation index values in 2020 and 2021 (Social Isolation Index), statewide monthly average income in 2020 (Income) in USD, and unemployment in 2020 (Unempl.).

| State | Region | Cases | Deaths | Auxílio 2020<br>Median (Min–Max) | Social Isolation Index (Median and 75% CI) |  | Income | Unempl. |
| --- | --- | --- | --- | --- | --- | --- | --- | --- |
|  |  |  |  |  | 2020 | 2021 |  |  |
| AC | N | 4653 | 88.9 | 27.8% (21.7%–30.5%) | 43.1% (38.1%–50.1%) | 44.9% (38.7%–52.1%) | \$167.55 | 15.1% |
| AM | N | 4777 | 126 | 28.4% (21.1%–30.7%) | 41.6% (36.6%–49.9%) | 42.1% (36.0%–47.5%) | \$155.68 | 15.8% |
| AP | N | 7914 | 107 | 30.0% (20.6%–32.6%) | 41.1% (36.8%–50.0%) | 48.5% (40.1%–55.5%) | \$163.17 | 14.9% |
| PA | N | 3378 | 82.7 | 28.3% (21.2%–30.8%) | 37.6% (33.5%–46.8%) | 37.0% (33.5%–49.7%) | \$161.34 | 10.4% |
| RO | N | 5329 | 101 | 25.9% (15.5%–28.9%) | 40.4% (36.3%–48.8%) | 39.6% (32.7%–47.1%) | \$213.60 | 10.4% |
| RR | N | 10883 | 124 | 29.0% (20.2%–31.8%) | 41.2% (36.6%–47.5%) | 43.1% (37.1%–51.1%) | \$179.61 | 16.4% |
| TO | N | 5682 | 77.6 | 25.4% (16.7%–27.4%) | 34.8% (31.5%–43.1%) | 33.9% (30.1%–43.0%) | \$193.68 | 11.6% |
| AL | NE | 3127 | 74.3 | 22.9% (17.6%–24.7%) | 39.4% (35.2%–45.8%) | 37.4% (33.7%–46.9%) | \$145.45 | 18.6% |
| BA | NE | 3305 | 61.1 | 25.1% (19.2%–26.8%) | 39.4% (35.7%–46.5%) | 38.0% (36.1%–48.0%) | \$176.32 | 19.8% |
| CE | NE | 3647 | 109 | 27.2% (20.7%–28.9%) | 40.9% (37.6%–48.6%) | 41.4% (39.7%–49.2%) | \$187.84 | 13.2% |
| MA | NE | 2824 | 63.3 | 26.2% (21.4%–28.6%) | 38.6% (35.7%–45.3%) | 39.7% (36.4%–45.3%) | \$123.52 | 15.9% |
| PB | NE | 4122 | 90.9 | 26.4% (20.9%–28.3%) | 39.6% (35.7%–45.1%) | 39.0% (35.4%–45.5%) | \$162.99 | 14.6% |
| PE | NE | 2310 | 100 | 23.9% (17.6%–25.3%) | 38.8% (35.1%–47.1%) | 41.2% (36.0%–47.1%) | \$163.90 | 16.8% |
| PI | NE | 4348 | 86.5 | 28.4% (22.6%–30.6%) | 39.6% (36.0%–48.3%) | 38.5% (33.2%–48.3%) | \$156.96 | 12.8% |
| RN | NE | 3341 | 84.7 | 26.1% (19.2%–27.7%) | 39.1% (34.3%–49.9%) | 35.9% (33.4%–49.9%) | \$ 196.79 | 15.8% |
| SE | NE | 4852 | 107 | 25.2% (19.0%–26.9%) | 38.3% (34.5%–44.0%) | 37.8% (34.0%–45.8%) | \$187.84 | 18.4% |
| DF | CW | 8239 | 139 | 19.1% (11.0%–21.9%) | 39.5% (35.9%–47.3%) | 38.2% (35.5%–48.2%) | \$452.23 | 14.8% |
| GO | CW | 4342 | 95.7 | 22.9% (13.6%–25.6%) | 36.1% (32.8%–43.7%) | 34.8% (32.5%–44.9%) | \$229.86 | 12.4% |
| MS | CW | 4761 | 82.9 | 22.6% (13.7%–25.4%) | 37.3% (34.2%–45.3%) | 37.0% (30.4%–44.1%) | \$271.89 | 10.0% |
| MT | CW | 5088 | 126 | 23.6% (14.0%–26.4%) | 37.2% (33.6%–45.5%) | 36.1% (30.8%–45.7%) | \$255.99 | 9.7% |
| ES | SE | 6108 | 125 | 24.4% (15.1%–27.2%) | 38.1% (33.8%–46.7%) | 36.8% (31.2%–46.9%) | \$246.12 | 12.7% |
| MG | SE | 2550 | 55.9 | 20.1% (12.8%–22.1%) | 38.0% (34.8%–45.1%) | 37.4% (31.1%–45.5%) | \$240.09 | 12.5% |
| RJ | SE | 2503 | 147 | 22.7% (14.4%–25.1%) | 39.0% (35.3%–44.8%) | 38.0% (35.0%–47.6%) | \$314.83 | 17.4% |
| SP | SE | 3159 | 101 | 17.7% (10.6%–20.0%) | 38.4% (34.1%–47.2%) | 36.2% (30.5%–46.1%) | \$331.45 | 13.9% |
| PR | S | 3617 | 69.2 | 19.8% (11.6%–22.3%) | 40.4% (37.1%–46.9%) | 39.3% (34.4%–47.0%) | \$275.54 | 9.4% |
| RS | S | 3937 | 77.7 | 17.8% (10.4%–20.6%) | 39.4% (34.3%–44.4%) | 41.5% (36.2%–48.2%) | \$321.40 | 9.1% |
| SC | S | 6792 | 72.4 | 16.3% (9.22%–19.3%) | 38.1% (33.6%–48.4%) | 37.5% (30.3%–48.6%) | \$298.20 | 6.1% |

**Figure 1:**
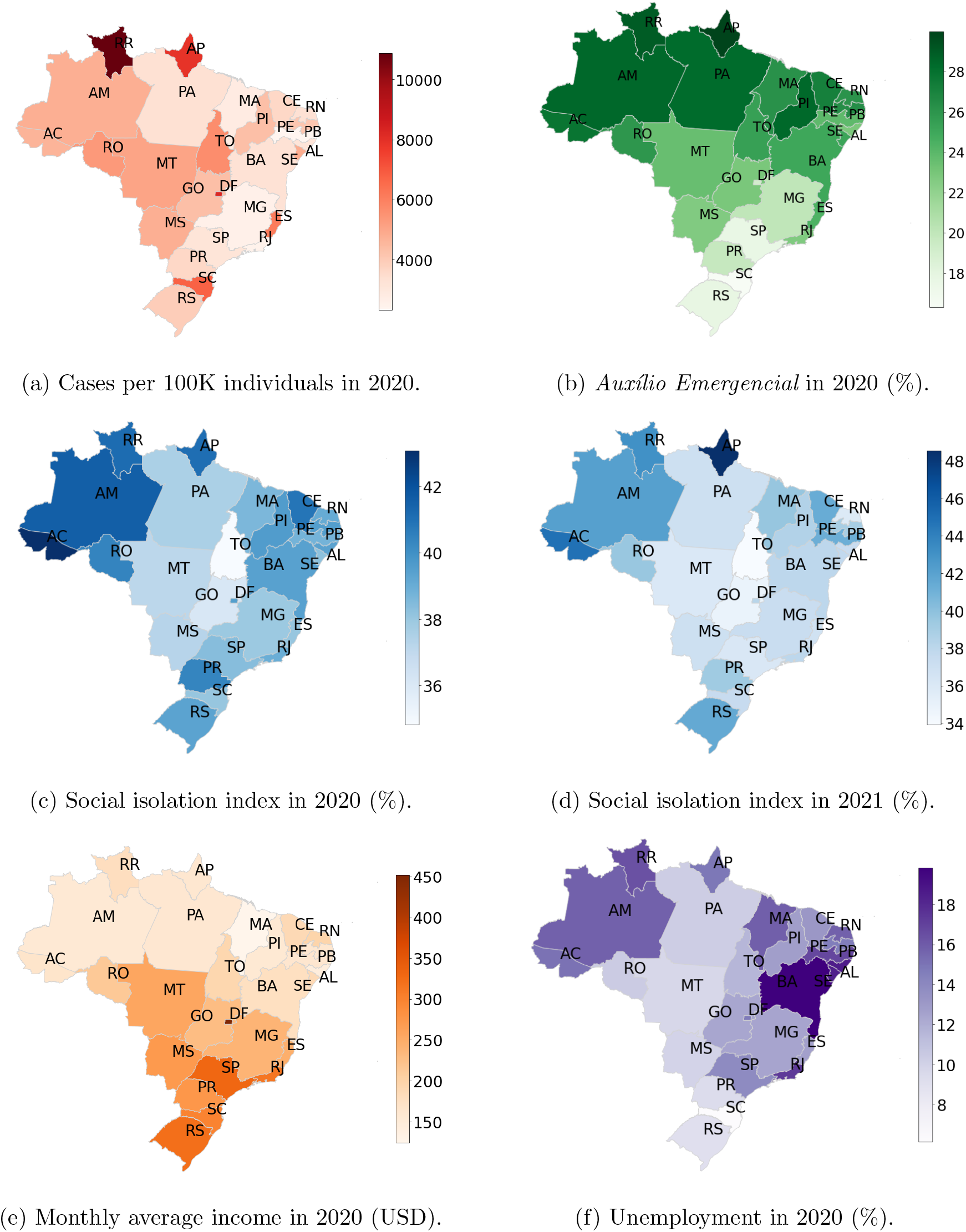
(a) Total number of COVID-19 infections per 100 thousand individuals for each state in Brazil during 2020. (b) Statewide median population proportions receiving the AE in 2020. Statewide median values of the social isolation index during 2020 (c) and 2021 (d). (e) Statewide monthly average income in 2020 (USD). (f) Statewide unemployment in 2020.

It is worth mentioning that, during the period of analysis, vacination in Brazil was not significant [41].

### Summary for the North Region

The North Region is composed of the states of Acre (AC), Amazonas (AM), Amapá (AP), Pará (PA), Rondônia (RO), Roraima (RR), and Tocantins (TO). In 2020, it concentrated 8.82% of the Brazilian population besides accounting for more than 45% of the Brazilian territory, which represented the smallest countrywide population density [42]. All the states in this region had, in 2020, a monthly average income smaller than the national average, and the unemployment higher than 10%. In the states of AC, AM, AP, and RR, the unemployment was higher than the national average of 13.5% [37, 38]. This indicates that a large proportion of the population in the region is socially and economically vulnerable. Such issues can have an important role in enhancing the impact of the AE as a social program and as a disease spread mitigation instrument, allowing individuals in the working age to stay at home.

As Figure 1 and Table 1 show, the North region proportionally received the largest amount of resources from the AE, since a median value of 28.1% (min–max 20.2%–30.2%) received a monthly income from April to December 2020, with monthly individual mean values of $143.91 USD during April to August and $87.12 BRL during September to December. In comparison with the monthly average income for each state, such payments represented from 63.7% to 91.8% and 45.2% to 55.2% in the first and the second periods, respectively. This drop in the monthly value seemed to impact negatively the capacity of individuals to adhere the social isolation, since, during such period, the social isolation index registered its lowest values for the majority of the states in the region, as the SII panels in Figures S.1–S.7 show.

Except for RR, all the states faced two large waves of infections, the first one starting on March or April 2020, and ending in August to October 2020. In general, the second wave started in January 2021, and, in some states, it was still ongoing in May 2021, as Figures S.1– S.7 illustrate. AE was suspended from January to March 2021, which means that, during the second wave, the states’ populations were economically vulnerable. This seems to have impacted social isolation since for the majority of the states in the North region, we could observe lower SII values during the second waves than in the first ones.

During the second wave, the health system in the State of Amazonas collapsed [43, 44], and a new COVID-19 strain (Gamma), which is potentially more contagious than the original one, was detected in the same state [45, 46, 47, 48]. Such issues were not sufficient to force the population in the region to adhere to social isolation at the same levels as the ones observed during the first waves. A number of reasons can be linked to such behavior, but the absence of the AE may have had an important impact, especially amongst those in the working age. One exception was RR that in January to March 2021 presented SII values similar to the ones observed during the first wave, which seems to have prevented a second wave of infections, as Figure S.6 illustrates.

The correlation between the daily variations in SII and in the reproduction number ℛ(*t*) was mainly negative, indicating that, an increase (or decrease) in SII is linked to a decrease (or increase) in ℛ(*t*). The median time between a change in SII to imply a change in ℛ(*t*) in the North region ranged from 14 days (50% CI: 12–17) in AC and 22 days (50% CI: 12–22) in AP. Correlation approached zero or became positive basically during periods when SII reached values below 40% or when SII and *ℛ* (*t*) stabilized around fixed values. The first case occurred mainly from August to December, when the payments from the AE were reduced. The second case occurred mainly during periods when the disease incidence was decreasing, ℛ(*t*) was close to 1.0, and SII was approximately 40% or larger. In these situations, it seems that SII loses its capacity of making ℛ(*t*) to change, especially if SII is small or large enough. As illustrated by Figure 1 and Table 1, the states with the largest population proportions receiving from the AE were AP and RR, which were exactly the ones that presented the largest regional numbers of COVID-19 infection per 100K individuals. Curiously, these states also presented high SII median values during the studied period. The large proportional number of COVID-19 infections is due to the first wave of infections since both states have managed to overcome or minimize the effects of a second wave of infections. In these two cases, the absence of the AE from January to March 2021 seems to have had little impact on the population adherence to social isolation.

Figure 1 and Table 1 also illustrate that AC, AM, AP, and RR had higher unemployment, lower statewide mean values of monthly income, larger population proportions receiving the AE in 2020, and larger SII median values. Thus, it seems that there is a relation between the AE, level of economical exposure, and adherence to social isolation in the following sense; amongst populations economically vulnerable, the AE has addressed such vulnerability for a population proportion sufficiently large to impact social isolation.

Therefore, the AE was of fundamental importance in providing economic support to families during the COVID-19 pandemic, which may have allowed a large proportion of the North Region population to stay at home, respecting social isolation measures.

### Summary for the Northeast Region

The Northeast Region is composed of the states of Alagoas (AL), Bahia (BA), Ceará (CE), Maranhão (MA), Paraíba (PB), Pernambuco (PE), Piauí (PI), Rio Grande do Norte (RN) and Sergipe (SE). In 2020, it concentrated 28% of the Brazilian population besides accounting for 18% of the Brazilian territory.

The statewide monthly average incomes varied from $123.52 USD (MA) to $196.79 USD (RN) in 2020. Thus, all the states in this region had in 2020, average incomes considerably lower than the national average income. Unemployment levels ranged from 12.8% (PI) to 18.58% (AL) in 2020. Except for PI and CE, all the states had unemployment levels higher than the national average, i.e., 13.5% [37, 38].

From May to November 2020, all the states in the Northeast region faced their first waves of infections, as Figs.S.8–S.16 show. The social isolation index presented small values, between 38.5% to 40.5% during 2020. It gradually decreased from April to October 2020, achieving values way below 40% in all states. From September to December 2020, there was a reduction of 44.0% in the amount paid by the AE. From October to January 2020, SII gradually increased, reaching approximately 40%. Then, it oscillated around to values below but close to 40% in March 2021. Such persistent low SII values probably contributed to the emergence of the second waves that started from November 2020 to January 2021. The second waves were still ongoing by the end of the period of analysis. For the major part of the states, the second waves coincided with the suspension of the AE.

During 2020, the states in this region registered between 2,310.2 (PE) and 4,851.8 (SE) infections per 100K individuals, and from January to May 2021 the accumulated cases represented already 36.8% (MA) and 89.4% (SE) of the registered infections in 2020.

From April to December 2020, the median proportion of individuals receiving the AE varied from 22.87% (AL) to 28.43% (PI). From April to August 2020, the mean monthly payments ranged from $151.91 USD (RN) to $180.48 USD (PE), representing 77.2% and 110.1% of the corresponding statewide average income. From September to December 2020, the monthly mean amount paid by the program dropped to values ranging from $82.81 USD (PI) to $103.46 USD (PE), representing 52.8% and 63.1% of the corresponding statewide average income. It is worth noticing that, PE and MA presented the lowest accumulated infections per 100K individuals in 2020.

Although presenting the second largest average income in the Northeast region, SE had, in 2020, the third largest unemployment, 18.4%. The SII in SE was mostly below 40% in 2020 and 25.2% on average of its population received the AE accordingly to Table S.1. Such persistent small SII values contributed to the larger accumulated infections per 100K individuals in 2020.

The correlation between the daily variations in SII and in the reproduction number ℛ(*t*) was mainly negative, indicating that, an increase (or decrease) in SII is linked to a decrease (or increase) in the reproduction number ℛ(*t*). The mean time between a change in SII to cause a change in ℛ(*t*) in the Northeast region ranged between 7 days (50% CI: 6–21) in SE and 25 days (50% CI: 16–30) in MA. During some periods, the correlation became close to zero or positive, especially in the periods from April to June, from August to October, and in December 2020. Such periods were also followed by elevated SII values, usually higher than 40%. Such SII values occurred mostly from the beginning of the period of analysis up to June or August 2020, depending on the state. Possibly the conjunction of the factors, such as ℛ(*t*) larger than one and SII lower than 40% resulted in zero or positive correlation.

### Summary for the Central-West Region

The Central-West Region has the second lowest regionwide population density. It is composed of the Distrito Federal (DF) and the states of Goiás (GO), Mato Grosso (MT), and Mato Grosso do Sul (MS). In 2020, the statewide monthly average income varied from $229.86 USD to $452.22 USD. Thus, except for Goiás, all the states had average incomes higher than the national average. Unemployment levels ranged from 9.73% to 14.75% in 2020. Apart from DF, all the states had a unemployment lower than the national average, i.e., 13.5% [37, 38].

From May to November 2020, all the states faced their first waves of infections, as shown in Figs. S.18–S.21. The social isolation index gradually decreased from April to October 2020, reaching values way below 40% in all the states. From October 2020 to January 2021, SII gradually increased, stabilizing around 40%. Then, it decreased again attaining values below 40% by March 2021. Such persistent low SII values possibly contributed to the emergence of second waves of infections that started in November 2020 in MS, in January 2021 in GO and MT, and in middle February 2021 in DF. The second waves were still ongoing by the end of the period of analysis.

As in other regions, the second waves coincided with the period of suspension of the AE. During 2020, the states of the Central-West region registered from 4,342.0 (GO) to 8,238.6 (DF) accumulated infections per 100K individuals, and from January to May 2021, the accumulated numbers varied from 84.54% to 108.46% of the registered infections in 2020.

From April to December 2020, the statewide median proportions of individuals receiving the AE varied from 19.1% to 23.6%. From April to August 2020, the mean of the monthly payments ranged from $139.53 USD (DF) to $145.82 USD (GO), then, from September to December 2020, the monthly mean amount dropped to the range from $99.05 USD (MS) to $105.67 USD (GO). Such values represent from 30.9% to 63.4% of the monthly statewide average income in the first period and from 23.2% to 46.0% in the second period.

The DF presented the largest SII values in 2020 in the region but they were mainly below 40%, which was apparently not enough to prevent a large number of accumulated infections per 100K individuals. Such difficulty in adhering to social isolation may be linked to a series of factors. Although DF is a high income region in comparison to other Brazilian states, the statewide rate of unemployment was also elevated in 2020. In addition, DF received the smallest amount of resources from the AE also in 2020. Possibly, the amount paid by the AE was not enough to support families, mainly because the income paid was small compared to the monthly average income in DF. This may have restricted the efficacy of the AE in keeping individuals at home, contributing to the observed high number of infections in this state.

The correlation between the daily variations in SII and in the reproduction number ℛ(*t*) was mainly negative, as expected. The median time between a change in SII to cause a change in ℛ(*t*) in the Central-West region ranged from 17 days (50% CI: 5–19) in GO to 19 days (50% CI: 14–27) in MS.

For the Center-West states, SII remained mostly below 40%. The loss of correlation between SII and ℛ(*t*) seems to occur in two situations, namely, when ℛ(*t*) is high, that is, far above one and SII is below 40 %, or when both ℛ(*t*) and SII are high. The second case occurred only at the beginning of the outbreak, when SII values were mostly above 40% and the estimated ℛ(*t*) was higher than one. The first case was more frequent especially between April to July 2020 and in December 2020. All the states in the Center-West region showed a loss of correlation in December.

The reduction in the values paid by the AE, from September to December 2020, and the program suspension from January to March 2021 apparently reduced the capacity of individuals to adhere to social isolation since, during such period, SII values were mainly lower than 40%. Moreover, when comparing SII values from April to June 2020 with those from January to March 2021, we noticed a non-negligible reduction, ranging from 17% to 6.3% in the monthly median values, which may have triggered or sustained second waves of infections.

### Summary for the Southeast Region

The Southeast Region has the largest countrywide population density. It is composed of the states of Espírito Santo (ES), Minas Gerais (MG), Rio de Janeiro (RJ), and São Paulo (SP). This was also the richest Brazilian region in 2020, with statewide monthly average incomes ranging from $240.09 USD in MG to $331.45 USD in SP. The 2020 unemployment levels in the region spanned from 12.5% in MG to 17.4% in RJ, meaning that large proportions of the state populations could be economically vulnerable. In 2020, the AE covered median proportions of 17.7% (min–max 10.6%–20.0%) in SP to 24.4% (min–max 15.1–27.2%) in ES, paying on average a monthly amount of $150.12 USD from April to August and $111.80 USD from September to December. The drop in the mean amount was 28.1%, and the mean payments represented proportions ranging from 48.5% to 63.3%, in the first period, and 34.1% to 44.0%, in the second one, of the statewide average incomes.

During the period of study, ES, MG and RJ faced three waves of infections, whereas SP faced two. The first waves were, in general, long lasting, initiating from March to May 2020 and ending in August to November 2020. Second waves, initiated in November 2020 and, in general, ended in February 2021. In SP the second wave was still ongoing by the end of the studied period, i.e., May 2021. Third waves began in February or March 2021 and were still ongoing by May 2021, i.e., the end of the study period. In the periods between waves of infections, daily reports remained large, representing non-negligible proportions of the peak of reports observed during the outbreaks.

The SII remained higher than 40% in the beginning of pandemic, mainly from March to July 2020, and from the end of December 2020 to middle January or middle February 2021, i.e., during the first and the second waves, respectively. SII values observed during the first waves are considerably larger than the ones observed in the second waves, representing a reduction from about 6% to approximately 20%, if we compare the median values of SII for April to June 2020, i.e., the first three months of operation of the AE, with those ones for January to March 2021, when the program was suspended. This may indicate a difficulty faced by proportions of individuals to adhere social isolation due to the economical exposure caused by the risk of unemployment and the absence of a program that could replace incomes during the lockdown. It is worth mentioning that, from August to December 2020, SII presented its smallest values, staying for long periods below 40%, which may have triggered the beginning of second waves and helped to keep the daily reports high during the periods between waves. In this period the monthly mean payments from the AE were reduced by 28.1%, which may have reduced the efficacy of the program in supporting those in need since the statewide monthly average incomes are considerably larger than the program payments.

Smaller SII values in 2021 may be linked to the large accumulated numbers of infections per 100K individuals from January to May 2021, since they already accounted for more than 81.8% in ES and 162.2% in MG of the registered infections in respective states in the whole 2020.

As expected, the correlation between the daily variations in SII and in the reproduction number ℛ(*t*) was mainly negative. The estimated median time between a change in SII to imply a change in ℛ(*t*) in the Southeast region ranged from 15 days (50% CI: 11–17) in ES to 20 days (50% CI: 20–22) in MG. Correlation approached zero or became positive basically during periods when SII reached values below 40% or when SII and ℛ(*t*) stabilized around fixed values. The first case occurred principally during August to December, when the payments from the AE were reduced. The second case occurred mainly during periods when the disease incidence was decreasing, ℛ(*t*) was close to 1.0, and SII was approximately 40% or larger.

Figure 1 and Table 1 show that the states with the largest population proportions receiving the AE were ES and RJ. ES and SP presented the largest regional numbers of COVID-19 infection per 100K individuals in 2020, whereas RJ presented one of the smallest. In 2020, RJ and SP presented the largest median SII values, whereas from January to March 2021, RJ and MG presented the largest median values. Although ES was the state with the largest coverage by the AE, it was the state with the smallest SII median values in 2020 and 2021, possibly due to the reduction in the monthly amount and the suspension of the social program, exposing those in need, since this is one of the poorest states in the Southeast region.

Figure 1 and Table 1 also illustrate that RJ had the largest unemployment level in 2020, the second largest coverage by the AE, and the largest median SII values in 2020, indicating a close relation between the AE and adherence to social isolation. SP also illustrates such relation, with smaller intensity, whereas in ES we could not identify such relation.

### Summary for the South Region

The South Region is composed of the states of Paraná (PR), Rio Grande do Sul (RS), and Santa Catarina (SC). In 2020, it concentrated 14.3% of the Brazilian population and accounted for approximately 6.80% of the Brazilian territory [42]. All the states in this region had, in 2020, monthly average incomes larger than the national average, ranging from $275.54 USD in PR to $321.40 USD in RS, and unemployment levels smaller than 10% spanning from 6.13% in SC to 9.38% in PR, which is considerably smaller than the countrywide rate, i.e., 13.5% [37, 38]. This means that, in comparison to other Brazilian regions, a smaller proportion of its population is economically and socially vulnerable, which leads us to expect a limited impact of the AE on influencing adherence to social isolation.

In 2020, South region proportionally received the smallest amount of resources from the AE, with median values of statewide population proportions ranging from 16.3% (min–max 9.22%–19.3%) in SC to 19.8% (min–max 11.6–22.3%) in PR, paying in mean a monthly amount of $141.15 USD during April to August and $106.66 USD during September to December. The drop in the mean amount was 24.4%, and the mean payments represented proportions of the statewide average incomes ranging from 43.9% to 51.8%, in the first period, and 32.8% to 38.3%, in the second one. In PR and RS, larger proportions of the population received the AE, larger median values of the social isolation index were observed, and smaller numbers of COVID-19 infections per 100K individuals were registered, when compared to SC, as Figure 1 and Table 1 show.

For all states, it was possible to observe three large waves of infections occurring almost simultaneously. The first waves occurred from April to October 2020, the second from November 2020 to January 2021, and the third from February to April 2021, as Figures S.26–S.28 illustrate. In general, the second and third waves were considerably larger than the first one, and, after larger outbreaks, the daily reports stabilized at numbers larger than the peak of the first wave. In other words, in this region, COVID-19 incidence was high, despite the reproduction number values stayed close to one during such periods.

The social isolation index presented similar values during the major outbreaks, regardless the presence of the AE, as expected, since the regional population is less economically vulnerable. States with larger proportions covered by the program presented larger SII values and smaller numbers of COVID-19 infections per 100K individuals in 2020. Moreover, SII remained higher than 40% at the beginning of the pandemic, mainly from March to July 2020, and from December 2020 to January or middle February 2021, i.e., during the first and the second waves, respectively. As in other regions, the SII values observed during the first waves were larger than the ones observed in the second waves, representing reductions from about 2% to approximately 19.5%, if we compare the median values of SII for April to June 2020, with those ones for January to March 2021. From August to December 2020, SII presented its smallest values, staying below 40%. Such period coincides with the one when the monthly mean payments from the AE were reduced by 24.4%.

As previously, smaller SII values in 2021 may be linked to the large accumulated numbers of infections at the beginning of 2021 (January to May), since they already accounted for more than 85.7% in SC and 138.3% in PR of the accumulated numbers per 100K individuals in respective states in the whole 2020.

Unemployment seemed to be linked to the AE coverage since for the states with a smaller unemployment level in 2020, smaller population proportions were covered by the program. On the other hand, monthly average income seemed to have little relation to the program coverage in the region, as we can observe in Figure 1 and Table 1.

The correlation between the daily variations in SII and in the reproduction number ℛ(*t*) was again mainly negative. The estimated median time between a change in SII to imply in a change in ℛ(*t*) in the South region was between 14 days (50% CI: 14–15) in PR and 28 days (50% CI: 28–28) in RS. As in other Brazilian regions, correlation approached zero or became positive basically during periods when SII reached values below 40% or when SII and ℛ(*t*) stabilized around fixed values. The first case generally occurred during the period from August to December, when the payments from the AE were reduced. The second case occurred basically during periods when the disease incidence was decreasing, ℛ(*t*) was close to 1.0, and SII was approximately 40% or larger.

### 3.1 The Impact of the AE on the Numbers of COVID-19 Cases and Deaths

During 2020, social isolation prevailed as one of the main diseases spread contention measures, helping to reduce the numbers of infections and deaths, as illustrated by the mainly negative values of the estimated correlation between the social isolation index and the reproduction number. The capability of adhering to social isolation is deeply affected by the individual’s economical and social status. Thus, social programs, like the AE, that provide income for those in need, help to allow individuals to stay at home, acting as a support to other disease contention measures.

By comparing the observed social isolation index values from April to August 2020 with those from January to March 2021, we could see remarkable reductions during the second period, reaching more than 20% in some states. This pattern can be related to several reasons including the AE, since, during the first period, the program was fully operational and in the second, it was suspended. Moreover, the first period includes the beginning of the first wave of infections in all states, whereas the second period includes the second or third waves.

To estimate the impact of the AE on controlling disease spread, we evaluated the potential accumulated COVID-19 infections from April to August 2020 if the social isolation index is reduced from 1.00 to 10.0 for each state. Such reduction values are related to the reduction observed when comparing SII values observed in the periods when the periods mentioned above, i.e, when the program was fully operational and when it was suspended. Table 2 and Figure 2 show the percentage of additional accumulated infections and deaths corresponding to the reductions in SII for the whole country. As the results show, the incremental numbers are considerably larger than the reports, illustrating the importance of social distancing and programs such as the AE to support disease spread control.

**Table 2:** Increment in the accumulated number of cases and deaths in the period from 01-April-2020 to 31-Aug-2020 if the Social Isolation Index is reduced in 1 to 10 points in Brazil. The numbers in the parentheses are 90% confidence intervals.

| Reduction | Cases (90% CI) | Deaths (90% CI) |
| --- | --- | --- |
| -1.00 | 58.7% (2.76%–369%) | 56.9% (2.50%–335%) |
| -2.00 | 74.5% (0.36%–421%) | 72.9% (0.92%–393%) |
| -3.00 | 39.8% (0.93%–279%) | 33.6% (1.07%–249%) |
| -4.00 | 21.8% (0.33%–190%) | 14.6% (0.24%–147%) |
| -5.00 | 10.2% (0.36%–259%) | 9.58% (0.22%–224%) |
| -6.00 | 44.7% (5.94%–480%) | 58.3% (9.29%–506%) |
| -7.00 | 58.5% (2.77%–793%) | 78.4% (4.22%–845%) |
| -8.00 | 232% (7.83%–1,223%) | 317% (0.42%–1,303%) |
| -9.00 | 309% (6.68%–1,623%) | 437% (10.8%–1,757%) |
| -10.0 | 651% (12.5%–2,131%) | 785% (14.6%–2,312%) |

**Figure 2:**
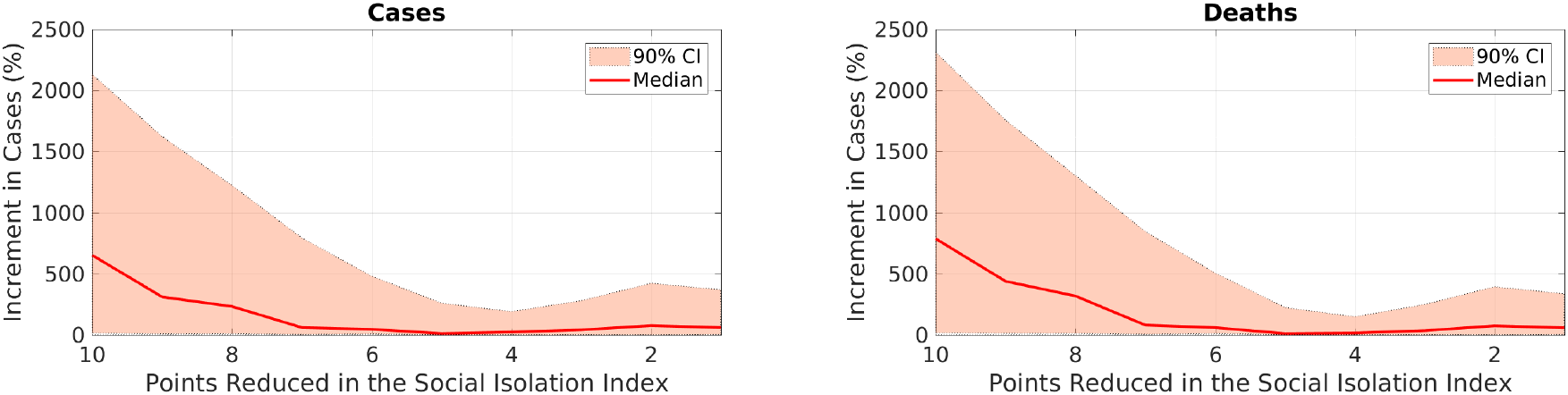
Increment in the accumulated number of cases and deaths in the period 01-April-2020 to 31-Aug-2020 if the Social Isolation Index is reduced in 1 to 10 points in Brazil. The dashed lines represent the 90% confidence interval.

## 4 Discussion

From March 2020 to May 2021, we observed two waves of COVID-19 infections in the majority of the states of the North, Northeast, and Central-West regions. On the other hand, almost all the States in the Southeast and South regions faced three waves of infection. The adherence to social isolation varied considerably across the country and time, but we observed similar patterns in the SII values. At the beginning of the first waves of infections, SII values were considerably higher than in other periods, even during secondary waves, where generally large numbers of infections were reported.

During the first waves, the AE was fully operational and reached a large fraction of the Brazilian population, paying a substantial proportion of the statewide average incomes. It probably helped individuals to stay at home, adhering to social isolation. Moreover, the period of reduction in the COVID-19 cases and the amount paid by the program generally coincided. It may have motivated people to leave social isolation, increasing SII values and triggering secondary waves of infections. From January to March 2021, when the program suspension occurred, second or third waves were ongoing in many states in the country. Maybe it may have made adherence to social isolation difficult, especially for those unemployed or precarious workers. In consequence, in this period, the SII values remained considerably lower than during the first wave.

The correlation between changes in the reproduction number and SII values was mainly negative in all states. Thus, if SII drops, we must expect that the reproduction number will rise. We also observed considerable delays between a rise in SII to cause a drop in the reproduction number. Moreover, such relation can also be lost when SII is low or the disease incidence is high.

In general, states with lower average income, in general, received proportionally more resources from the AE and presented higher SII values and lower numbers of accumulated infections per 100K individuals.

Based on the differences between the SII values observed in the first waves and those from January to March 2021, we estimated the accumulated numbers of infections and deaths from April to August 2020 if SII is reduced from 1 to 10 points. Based on these estimates, we found that the AE probably avoided a considerable number of infections and deaths during the referred period, when the program was fully operational.

It is worth mentioning that the proposed methodology has some limitations. We cannot assert that the presence or the absence of the AE was the main factor in promoting adherence to social isolation. Other factors, such as the fear of an emerging deadly disease, may have helped to convince people to stay at home at the beginning of the outbreak. Also, psychological saturation can be one of the main reasons why people leave isolation. These factors are hard to include in a model and can affect the cause and effect relationship between socioeconomic programs and social isolation, as well as the disease spread. More sophisticated statistical tools can be used to investigate further such causal relationships, and they are the subject of future work.

Different measures of mobility mainly based on mobile phone information were used to infer the real impact of initiatives like lockdowns on the contention of outbreaks [22, 24, 25, 26], as well as to describe the Spatio-temporal dynamics of the disease [19, 20, 23, 27, 28]. In the present work, we intended to shed light on the impact of a national socioeconomic program on disease spread contention, based on the premise that social isolation implies disease spread contention. As mentioned above, such a premise was widely tested and illustrated by our results.

## 5 Concluding Remarks

In 2020, the AE covered a median proportion of 22.1% (min–max: 14.7%–24.1%) of the Brazilian population providing income for those in need, under a nationwide unemployment of 13.5%. Comparing the SII values for the period when the socioeconomic program was fully operational with the period when it was not, we observe a significantly higher adherence to social isolation in the first period for the majority of the Brazilian states. It made us believe that the AE played a significant role in helping people to adhere to social isolation, helping to reduce the virus spread. Of course, there are other possible reasons for such differences in social isolation patterns since, during secondary waves, COVID-19 was not a novelty anymore. However, even during the collapse of the health system in Manaus in January 2021 and the introduction of a new virus strain potentially more contagious, the SII values were considerably smaller during the second or third waves in all Brazilian states.

We used such differences in SII values to estimate the impact of the absence of the AE, and concluded that the program potentially avoided a considerable number of COVID-19 infections and related deaths. The socioeconomic program provided income to a substantial proportion of the Brazilian population, including unemployed individuals and precarious workers, allowing them to stay at home. Without such resources, they would be gathering and helping the virus to spread. When the program was operational was sufficient to give time for public authorities to prepare the Brazilian health system to treat those with severe or critical symptoms.

Thus, if non-pharmaceutical measures, like lockdowns, are the only available ways to control an outbreak, socioeconomic programs can play an important supporting role by providing conditions to proportions of the population to adhere to social isolation. Coordinating such programs with sufficiently elevated coverage during a health crisis is a daunting task, but it can be crucial to make disease spread control succeed.

## Data Availability

All data produced in the present work are contained in the manuscript and available at a GitHub repository.

https://github.com/viniciusalbani/AuxilioEmergencial

## Data Availability

The datasets analyzed and used during the present study are available in [5, 7, 29, 36, 37, 38].

## Code Availability

The codes in MATLAB (The MathWorks, Inc., Natick, USA) used in this study can be found in the GitHub repository https://github.com/viniciusalbani/AuxilioEmergencial.

## Acknowledgments

VA acknowledges the financial support from Fundação Butantan through the Grant 01/2020. RA acknowledges the financial support from Fundação Carlos Chagas Filho de Amparo à Pesquisa do Estado do Rio de Janeiro (FAPERJ) through the grants E-26/202.932/2019 and E-26/202.933/2019. EM acknowledges the financial support from Conselho Nacional de Desenvolvimento Científico e Tecnológico (CNPq) and Fundação Butantan through the Grants 305544/2011-0 and 01/2020, respectively. JZ acknowledges the financial support from Khalifa University, CNPq, and Fundação Carlos Chagas Filho de Amparo à Pesquisa do Estado do Rio de Janeiro through the Grants FSU-2020-09, 307873/2013-7, and E-26/202.927/2017, respectively. All the authors thank Edilson Moura from Icognia (incognia.com) for sharing the Social Isolation Index dataset.

## Author contributions statement

V.A., E.M., and J.Z. designed research; V.A. and R.A. performed the research; V.A., R.A., and N.B. analyzed data; all the authors wrote the paper.

