## supplement for "On the Role of Financial Support Programs in Mitigating the Sars-CoV-2 Spread in Brazil"

November 27, 2021

[1]Department of Mathematics, Federal University of Santa Catarina, Florianopolis, Brazil,
[2]Instituto Politecnico do Rio de Janeiro, State University of Rio de Janeiro, Nova Friburgo, Brazil,
[3]Federal University of Technology - Paraná, Curitiba, Brazil,
[4]School of Medicine, University of São Paulo and LIM01-HCFMUSP, São Paulo, Brazil
[5]School of Applied Mathematics, Fundação Getúlio Vargas, Rio de Janeiro, Brazil,
[6]Mathematics Department, Khalifa University, Abu Dhabi, UAE,

### 1 North Region

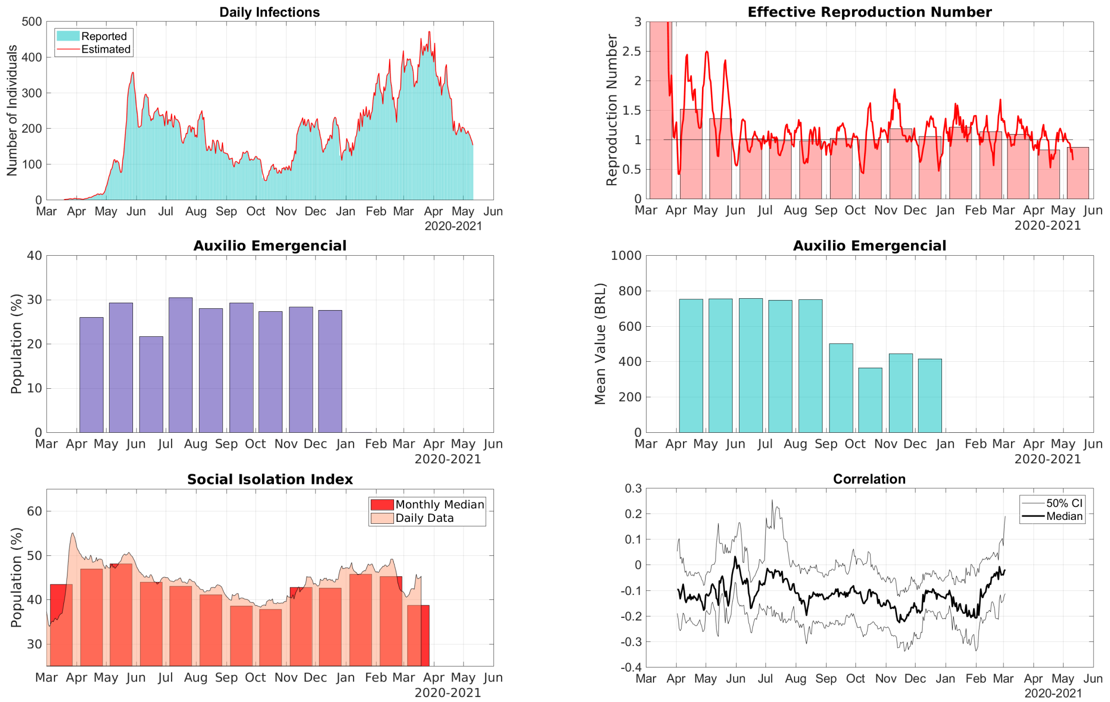


Figure S.1: Data for the State of Acre in the North Region of Brazil. Top Row, Right: comparison between the 7-day moving average of daily reports of infections and model predictions (right). Top Row, Left: The solid line represents the time-dependent reproduction number and the bars are the corresponding monthly median values. Middle Row, Right: The bars represent the proportions of the State population receiving the Auxílio Emergencial by month. Middle Row, Left: The bars represent the statewide average amount paid by Auxílio Emergencial each month. Bottom Row, Left: The area graph is the 7-day moving average of the social isolation index and the bars represent the corresponding monthly median values. Bottom Row, Right: Correlation between the daily increments of the social isolation index and the reproduction number.

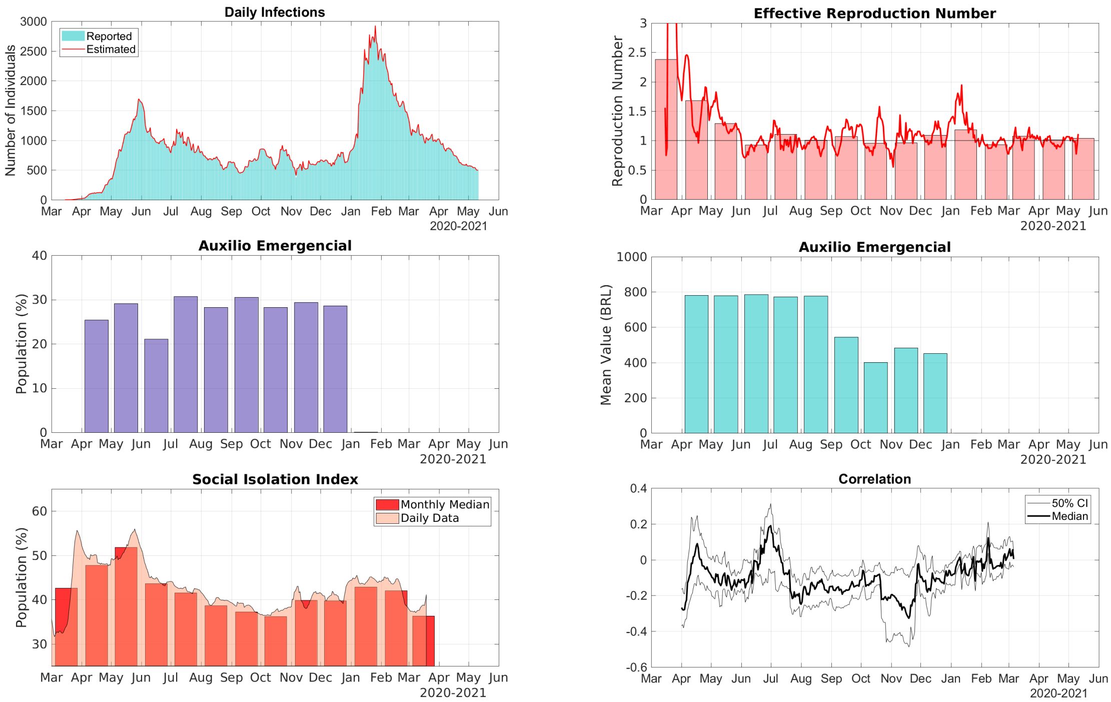


Figure S.2: Data for the State of Amazonas in the North Region of Brazil. Top Row, Right: comparison between the 7-day moving average of daily reports of infections and model predictions (right). Top Row, Left: The solid line represents the time-dependent reproduction number and the bars are the corresponding monthly median values. Middle Row, Right: The bars represent the proportions of the State population receiving the Auxílio Emergencial by month. Middle Row, Left: The bars represent the statewide average amount paid by Auxílio Emergencial each month. Bottom Row, Left: The area graph is the 7-day moving average of the social isolation index and the bars represent the corresponding monthly median values. Bottom Row, Right: Correlation between the daily increments of the social isolation index and the reproduction number.

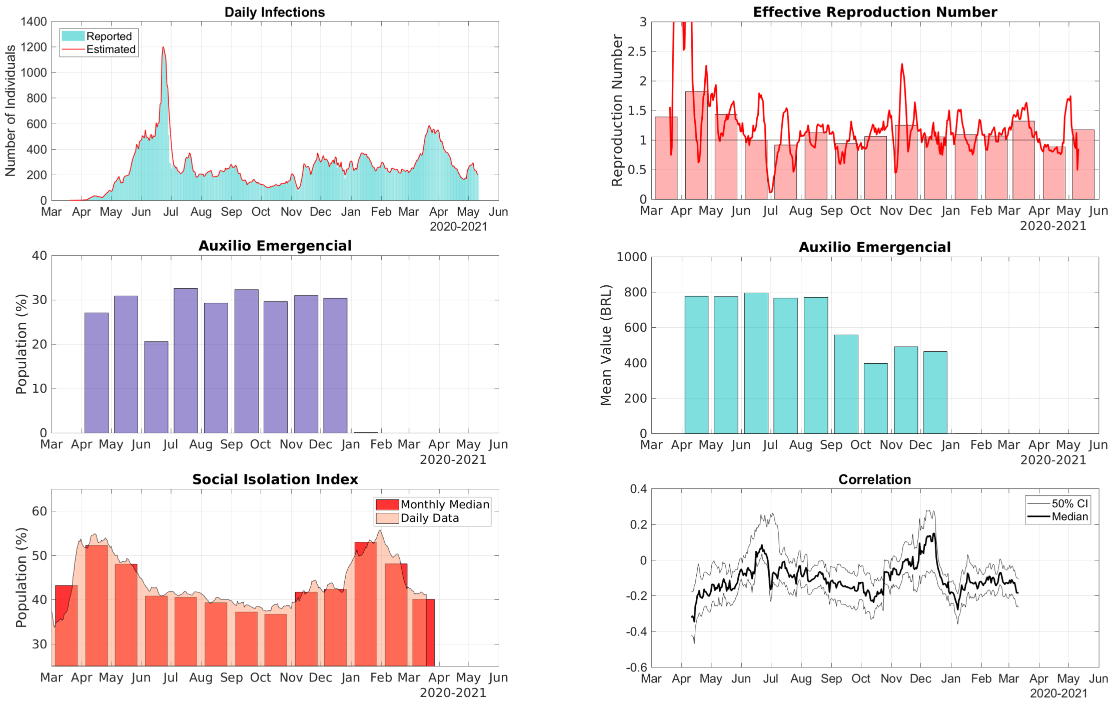


Figure S.3: Data for the State of Amapá in the North Region of Brazil. Top Row, Right: comparison between the 7-day moving average of daily reports of infections and model predictions (right). Top Row, Left: The solid line represents the time-dependent reproduction number and the bars are the corresponding monthly median values. Middle Row, Right: The bars represent the proportions of the State population receiving the Auxílio Emergencial by month. Middle Row, Left: The bars represent the statewide average amount paid by Auxílio Emergencial each month. Bottom Row, Left: The area graph is the 7-day moving average of the social isolation index and the bars represent the corresponding monthly median values. Bottom Row, Right: Correlation between the daily increments of the social isolation index and the reproduction number.

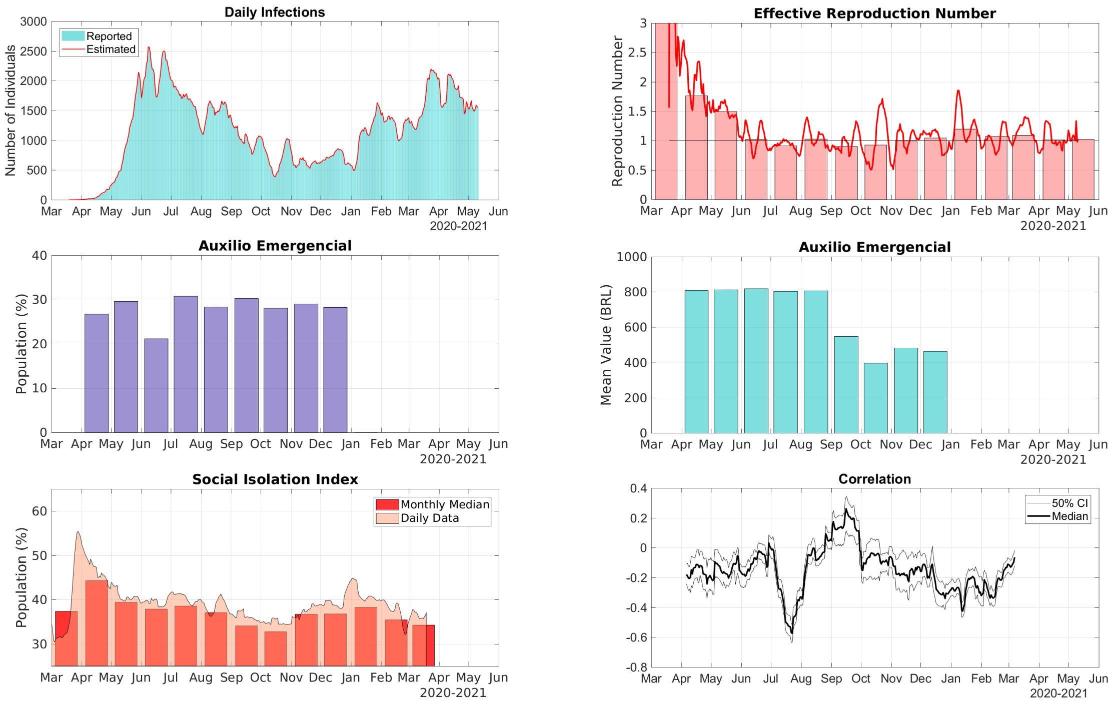


Figure S.4: Data for the State of Pará in the North Region of Brazil. Top Row, Right: comparison between the 7-day moving average of daily reports of infections and model predictions (right). Top Row, Left: The solid line represents the time-dependent reproduction number and the bars are the corresponding monthly median values. Middle Row, Right: The bars represent the proportions of the State population receiving the Auxílio Emergencial by month. Middle Row, Left: The bars represent the statewide average amount paid by Auxílio Emergencial each month. Bottom Row, Left: The area graph is the 7-day moving average of the social isolation index and the bars represent the corresponding monthly median values. Bottom Row, Right: Correlation between the daily increments of the social isolation index and the reproduction number.

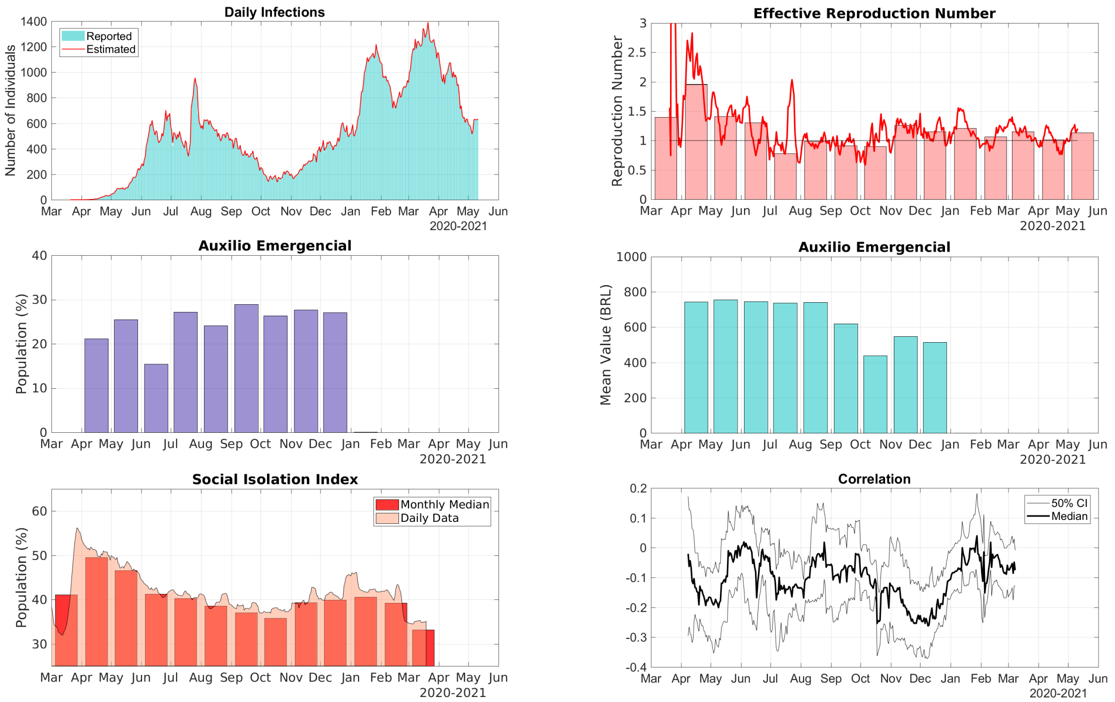


Figure S.5: Data for the State of Rondônia in the North Region of Brazil. Top Row, Right: comparison between the 7-day moving average of daily reports of infections and model predictions (right). Top Row, Left: The solid line represents the time-dependent reproduction number and the bars are the corresponding monthly median values. Middle Row, Right: The bars represent the proportions of the State population receiving the Auxílio Emergencial by month. Middle Row, Left: The bars represent the statewide average amount paid by Auxílio Emergencial each month. Bottom Row, Left: The area graph is the 7-day moving average of the social isolation index and the bars represent the corresponding monthly median values. Bottom Row, Right: Correlation between the daily increments of the social isolation index and the reproduction number.

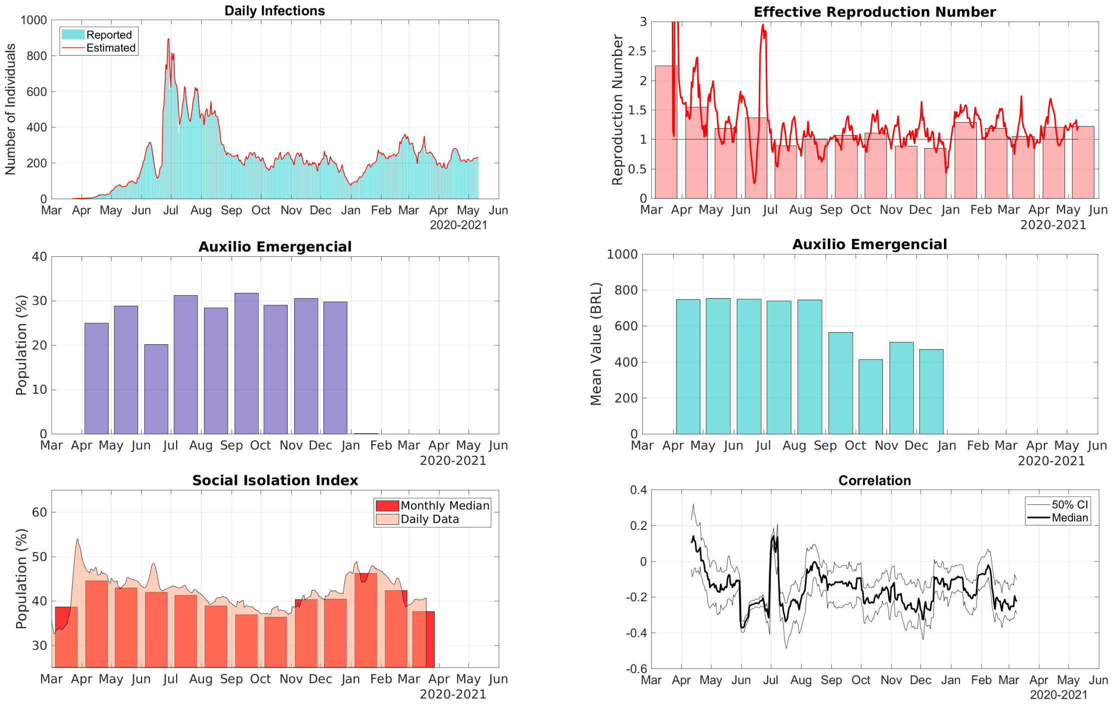


Figure S.6: Data for the State of Roraima in the North Region of Brazil. Top Row, Right: comparison between the 7-day moving average of daily reports of infections and model predictions (right). Top Row, Left: The solid line represents the time-dependent reproduction number and the bars are the corresponding monthly median values. Middle Row, Right: The bars represent the proportions of the State population receiving the Auxílio Emergencial by month. Middle Row, Left: The bars represent the statewide average amount paid by Auxílio Emergencial each month. Bottom Row, Left: The area graph is the 7-day moving average of the social isolation index and the bars represent the corresponding monthly median values. Bottom Row, Right: Correlation between the daily increments of the social isolation index and the reproduction number.

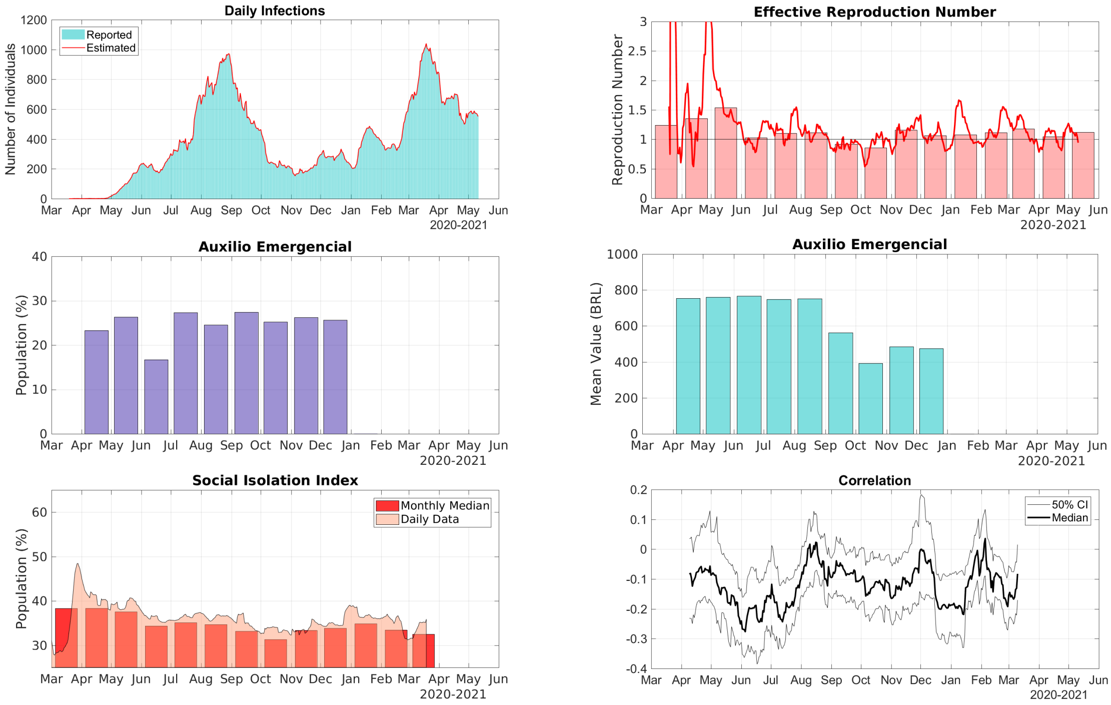


Figure S.7: Data for the State of Tocantins in the North Region of Brazil. Top Row, Right: comparison between the 7-day moving average of daily reports of infections and model predictions (right). Top Row, Left: The solid line represents the time-dependent reproduction number and the bars are the corresponding monthly median values. Middle Row, Right: The bars represent the proportions of the State population receiving the Auxílio Emergencial by month. Middle Row, Left: The bars represent the statewide average amount paid by Auxílio Emergencial each month. Bottom Row, Left: The area graph is the 7-day moving average of the social isolation index and the bars represent the corresponding monthly median values. Bottom Row, Right: Correlation between the daily increments of the social isolation index and the reproduction number.

### 2 Northeast Region

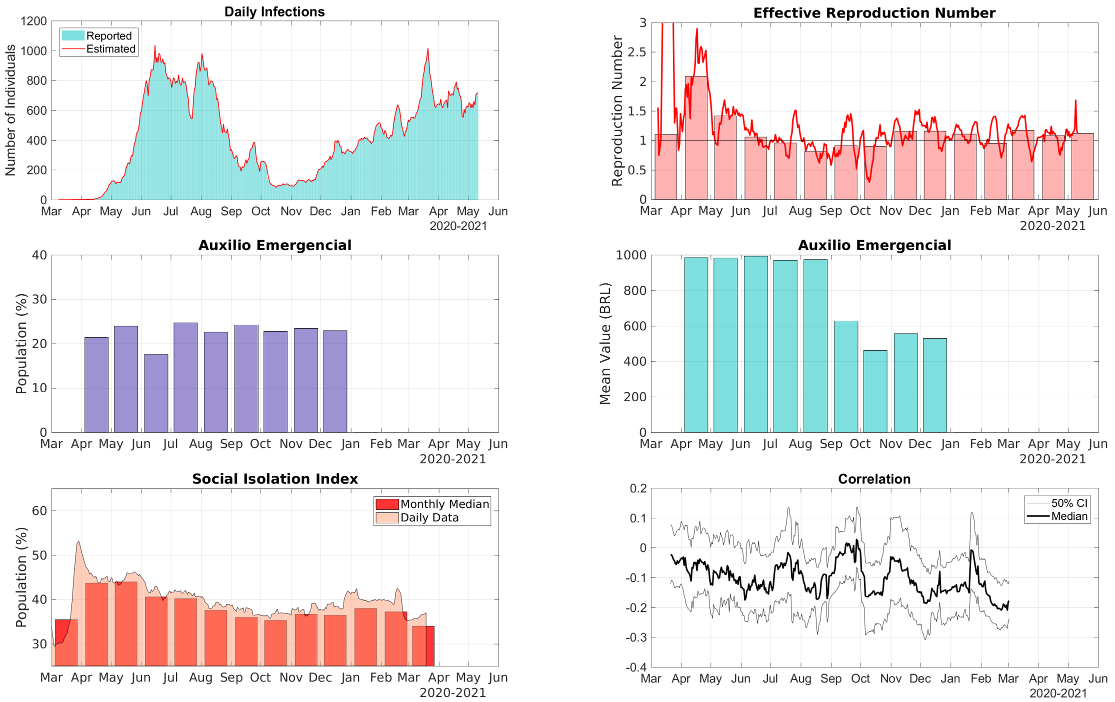


Figure S.8: Data for the State of Alagoas in the Northeast Region of Brazil. Top Row, Right: comparison between the 7-day moving average of daily reports of infections and model predictions (right). Top Row, Left: The solid line represents the time-dependent reproduction number and the bars are the corresponding monthly median values. Middle Row, Right: The bars represent the proportions of the State population receiving the Auxílio Emergencial by month. Middle Row, Left: The bars represent the statewide average amount paid by Auxílio Emergencial each month. Bottom Row, Left: The area graph is the 7-day moving average of the social isolation index and the bars represent the corresponding monthly median values. Bottom Row, Right: Correlation between the daily increments of the social isolation index and the reproduction number.

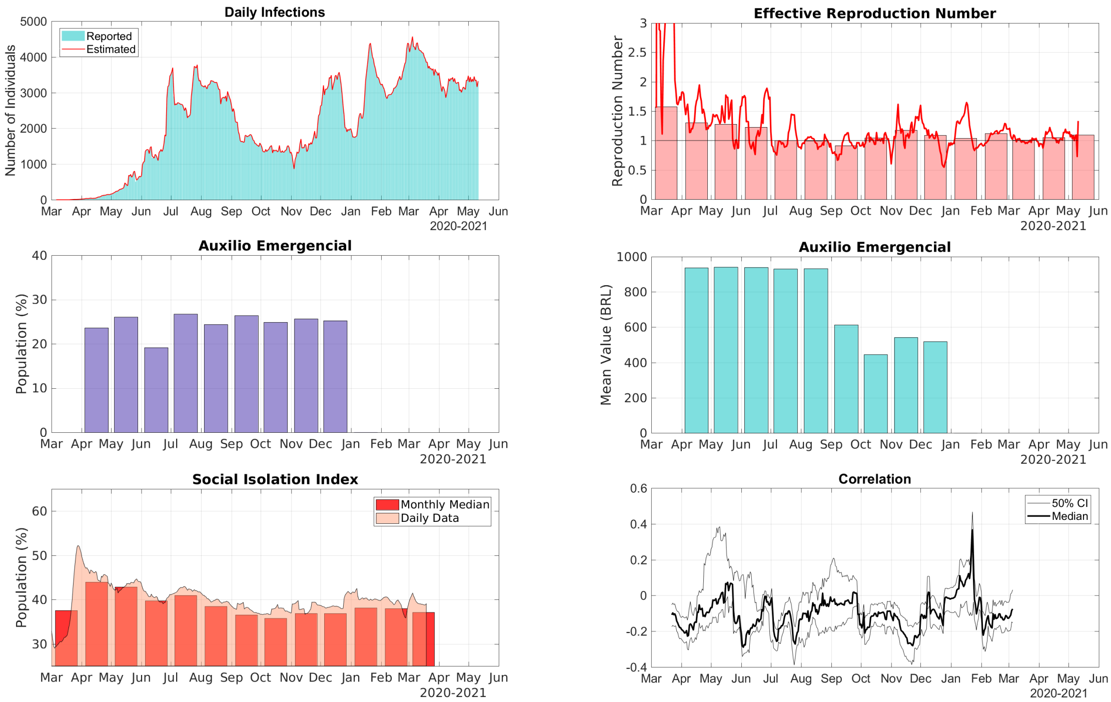


Figure S.9: Data for the State of Bahia in the Northeast Region of Brazil. Top Row, Right: comparison between the 7-day moving average of daily reports of infections and model predictions (right). Top Row, Left: The solid line represents the time-dependent reproduction number and the bars are the corresponding monthly median values. Middle Row, Right: The bars represent the proportions of the State population receiving the Auxílio Emergencial by month. Middle Row, Left: The bars represent the statewide average amount paid by Auxílio Emergencial each month. Bottom Row, Left: The area graph is the 7-day moving average of the social isolation index and the bars represent the corresponding monthly median values. Bottom Row, Right: Correlation between the daily increments of the social isolation index and the reproduction number.

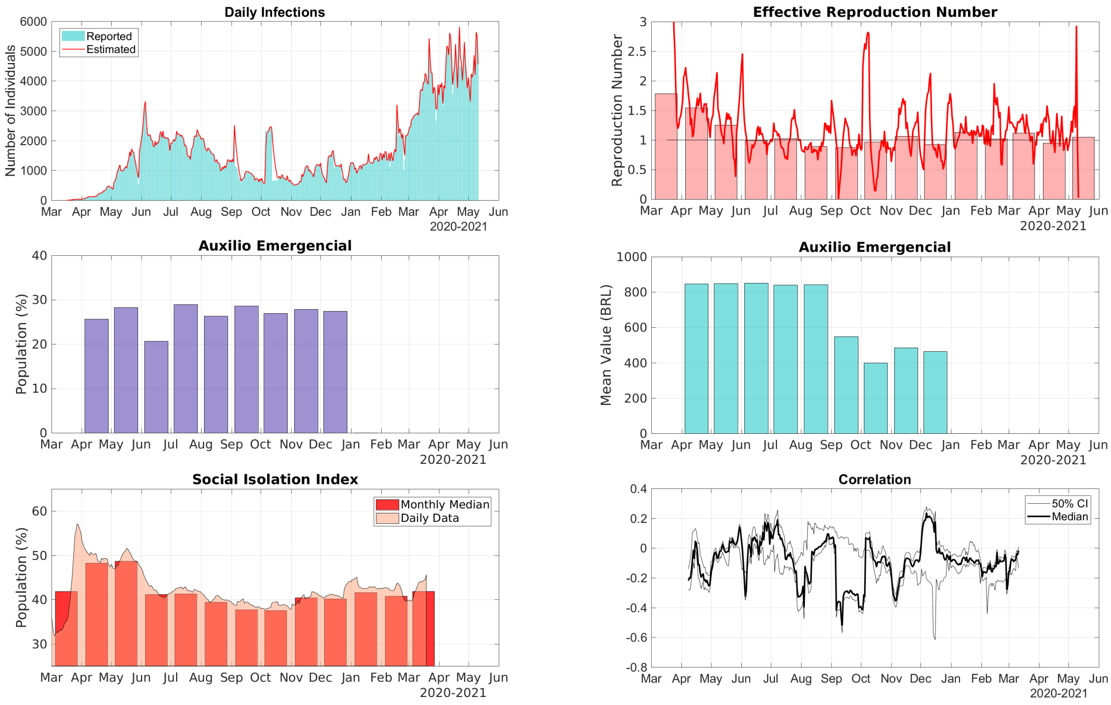


Figure S.10: Data for the State of Ceará in the Northeast Region of Brazil. Top Row, Right: comparison between the 7-day moving average of daily reports of infections and model predictions (right). Top Row, Left: The solid line represents the time-dependent reproduction number and the bars are the corresponding monthly median values. Middle Row, Right: The bars represent the proportions of the State population receiving the Auxílio Emergencial by month. Middle Row, Left: The bars represent the statewide average amount paid by Auxílio Emergencial each month. Bottom Row, Left: The area graph is the 7-day moving average of the social isolation index and the bars represent the corresponding monthly median values. Bottom Row, Right: Correlation between the daily increments of the social isolation index and the reproduction number.

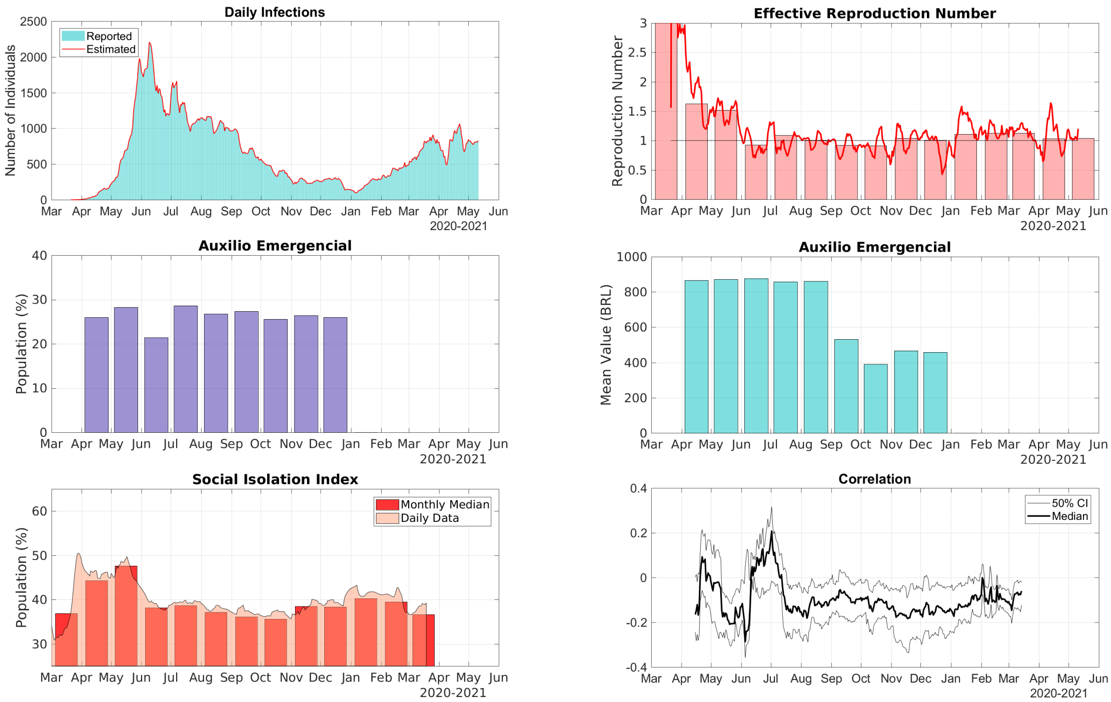


Figure S.11: Data for the State of Maranhão in the Northeast Region of Brazil. Top Row, Right: comparison between the 7-day moving average of daily reports of infections and model predictions (right). Top Row, Left: The solid line represents the time-dependent reproduction number and the bars are the corresponding monthly median values. Middle Row, Right: The bars represent the proportions of the State population receiving the Auxílio Emergencial by month. Middle Row, Left: The bars represent the statewide average amount paid by Auxílio Emergencial each month. Bottom Row, Left: The area graph is the 7-day moving average of the social isolation index and the bars represent the corresponding monthly median values. Bottom Row, Right: Correlation between the daily increments of the social isolation index and the reproduction number.

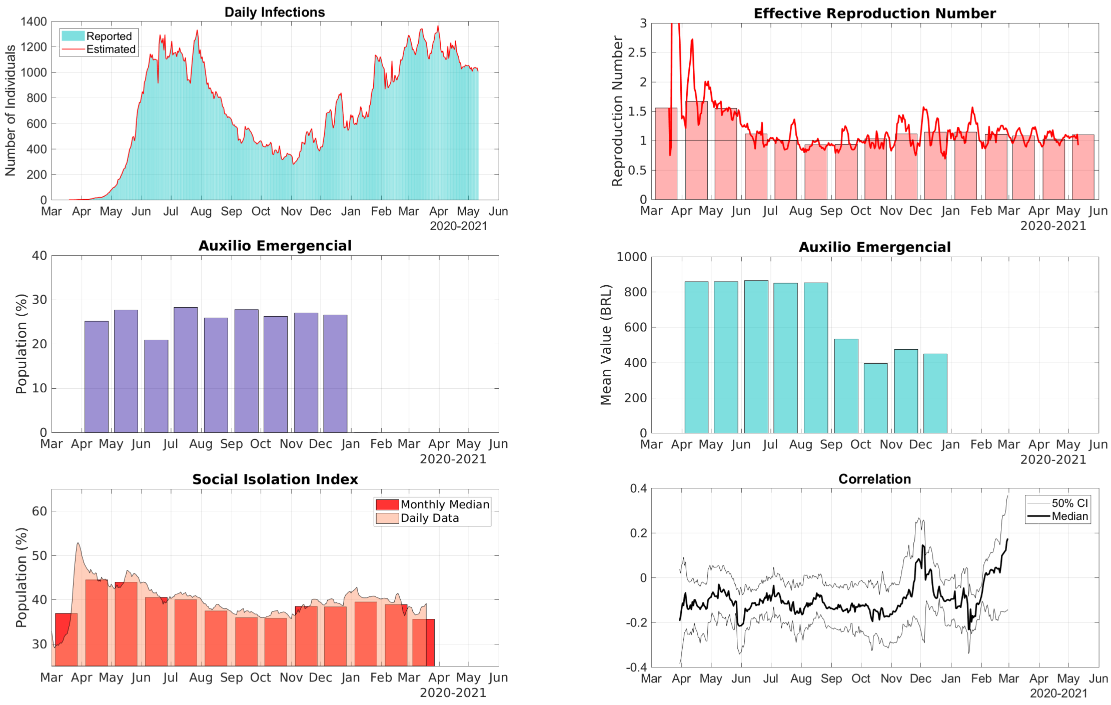


Figure S.12: Data for the State of Paraíba in the Northeast Region of Brazil. Top Row, Right: comparison between the 7-day moving average of daily reports of infections and model predictions (right). Top Row, Left: The solid line represents the time-dependent reproduction number and the bars are the corresponding monthly median values. Middle Row, Right: The bars represent the proportions of the State population receiving the Auxílio Emergencial by month. Middle Row, Left: The bars represent the statewide average amount paid by Auxílio Emergencial each month. Bottom Row, Left: The area graph is the 7-day moving average of the social isolation index and the bars represent the corresponding monthly median values. Bottom Row, Right: Correlation between the daily increments of the social isolation index and the reproduction number.

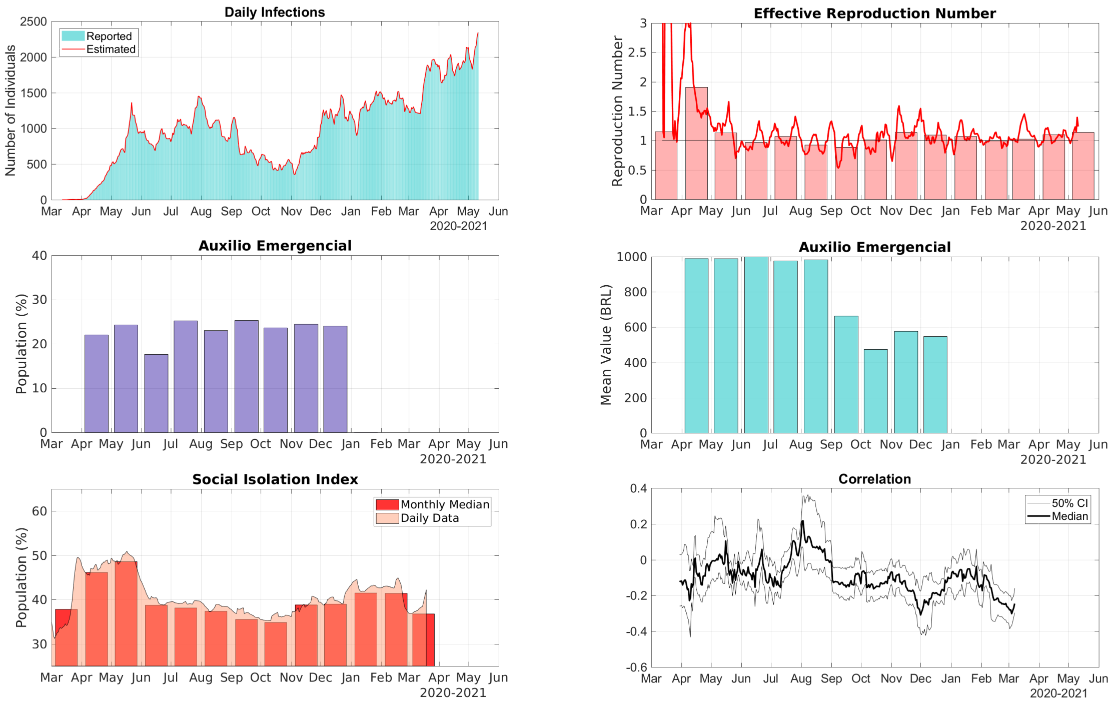


Figure S.13: Data for the State of Pernambuco in the Northeast Region of Brazil. Top Row, Right: comparison between the 7-day moving average of daily reports of infections and model predictions (right). Top Row, Left: The solid line represents the time-dependent reproduction number and the bars are the corresponding monthly median values. Middle Row, Right: The bars represent the proportions of the State population receiving the Auxílio Emergencial by month. Middle Row, Left: The bars represent the statewide average amount paid by Auxílio Emergencial each month. Bottom Row, Left: The area graph is the 7-day moving average of the social isolation index and the bars represent the corresponding monthly median values. Bottom Row, Right: Correlation between the daily increments of the social isolation index and the reproduction number.

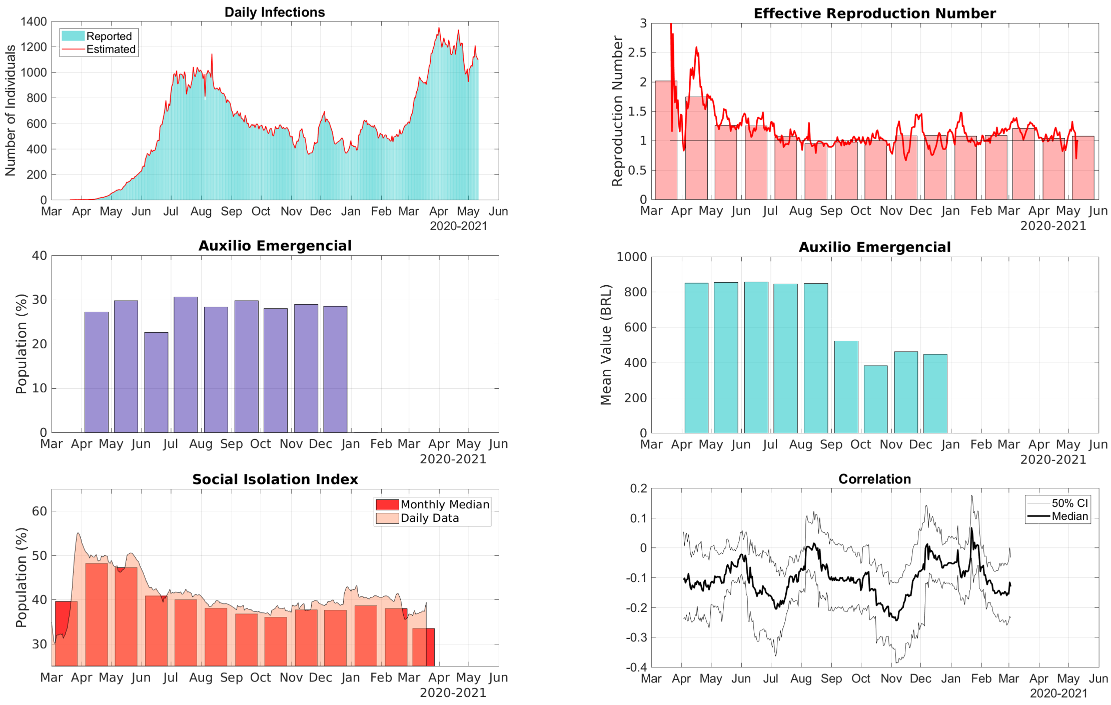


Figure S.14: Data for the State of Piauí in the Northeast Region of Brazil. Top Row, Right: comparison between the 7-day moving average of daily reports of infections and model predictions (right). Top Row, Left: The solid line represents the time-dependent reproduction number and the bars are the corresponding monthly median values. Middle Row, Right: The bars represent the proportions of the State population receiving the Auxílio Emergencial by month. Middle Row, Left: The bars represent the statewide average amount paid by Auxílio Emergencial each month. Bottom Row, Left: The area graph is the 7-day moving average of the social isolation index and the bars represent the corresponding monthly median values. Bottom Row, Right: Correlation between the daily increments of the social isolation index and the reproduction number.

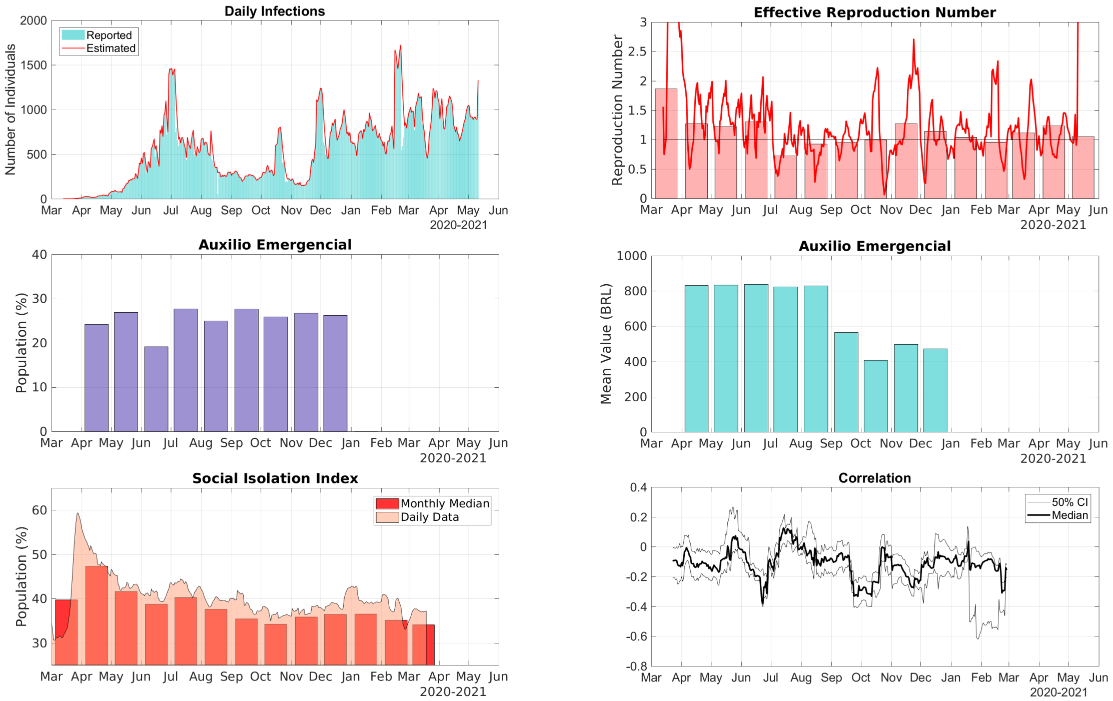


Figure S.15: Data for the State of Rio Grande do Norte in the Northeast Region of Brazil. Top Row, Right: comparison between the 7-day moving average of daily reports of infections and model predictions (right). Top Row, Left: The solid line represents the time-dependent reproduction number and the bars are the corresponding monthly median values. Middle Row, Right: The bars represent the proportions of the State population receiving the Auxílio Emergencial by month. Middle Row, Left: The bars represent the statewide average amount paid by Auxílio Emergencial each month. Bottom Row, Left: The area graph is the 7-day moving average of the social isolation index and the bars represent the corresponding monthly median values. Bottom Row, Right: Correlation between the daily increments of the social isolation index and the reproduction number.

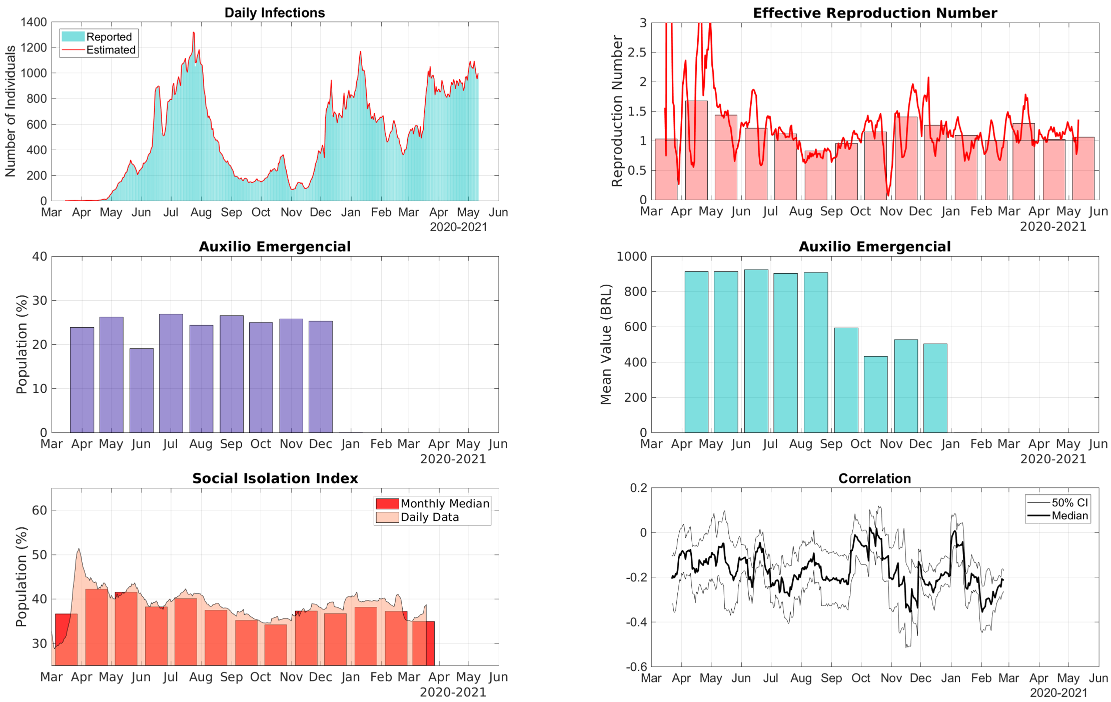


Figure S.16: Data for the State of Sergipe in the Northeast Region of Brazil. Top Row, Right: comparison between the 7-day moving average of daily reports of infections and model predictions (right). Top Row, Left: The solid line represents the time-dependent reproduction number and the bars are the corresponding monthly median values. Middle Row, Right: The bars represent the proportions of the State population receiving the Auxílio Emergencial by month. Middle Row, Left: The bars represent the statewide average amount paid by Auxílio Emergencial each month. Bottom Row, Left: The area graph is the 7-day moving average of the social isolation index and the bars represent the corresponding monthly median values. Bottom Row, Right: Correlation between the daily increments of the social isolation index and the reproduction number.

### 3 Central-West Region

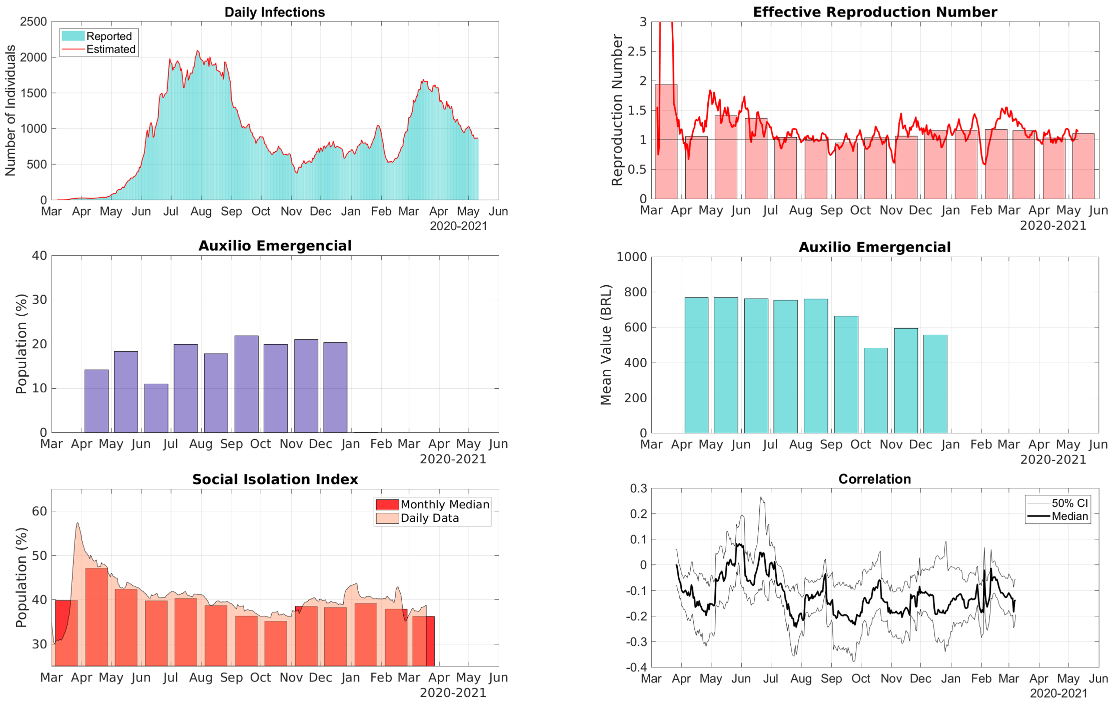


Figure S.17: Data for the Distrito Federal in the Central-West Region of Brazil. Top Row, Right: comparison between the 7-day moving average of daily reports of infections and model predictions (right). Top Row, Left: The solid line represents the time-dependent reproduction number and the bars are the corresponding monthly median values. Middle Row, Right: The bars represent the proportions of the State population receiving the Auxílio Emergencial by month. Middle Row, Left: The bars represent the statewide average amount paid by Auxílio Emergencial each month. Bottom Row, Left: The area graph is the 7-day moving average of the social isolation index and the bars represent the corresponding monthly median values. Bottom Row, Right: Correlation between the daily increments of the social isolation index and the reproduction number.

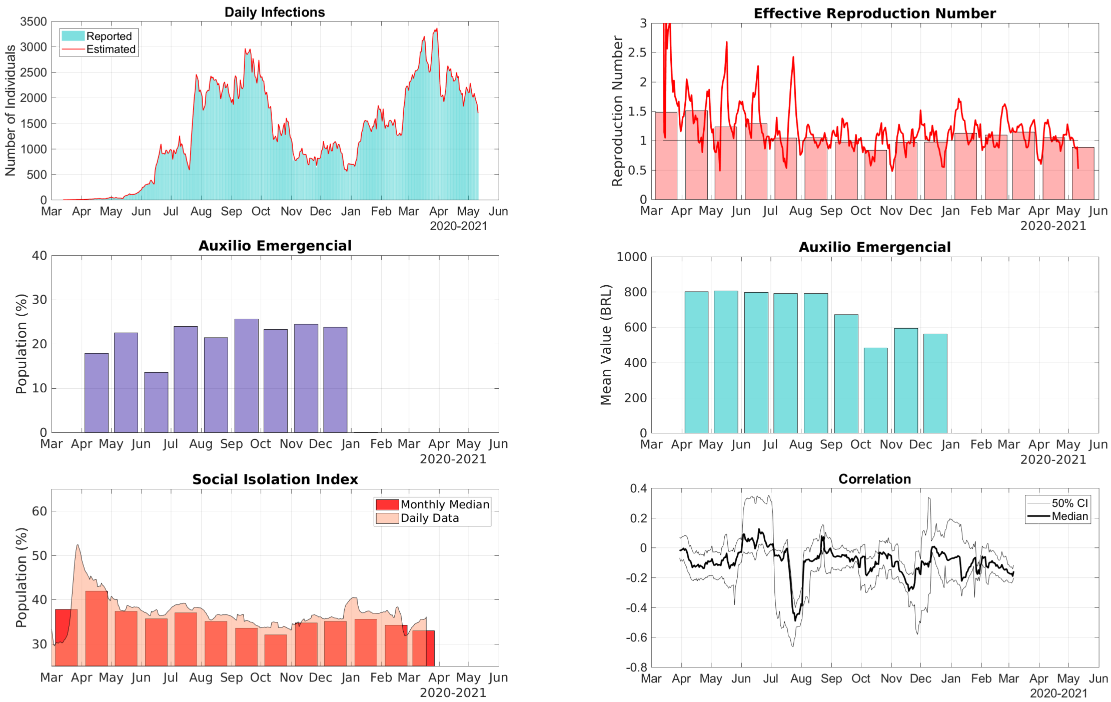


Figure S.18: Data for the State of Goiás in the Central-West Region of Brazil. Top Row, Right: comparison between the 7-day moving average of daily reports of infections and model predictions (right). Top Row, Left: The solid line represents the time-dependent reproduction number and the bars are the corresponding monthly median values. Middle Row, Right: The bars represent the proportions of the State population receiving the Auxílio Emergencial by month. Middle Row, Left: The bars represent the statewide average amount paid by Auxílio Emergencial each month. Bottom Row, Left: The area graph is the 7-day moving average of the social isolation index and the bars represent the corresponding monthly median values. Bottom Row, Right: Correlation between the daily increments of the social isolation index and the reproduction number.

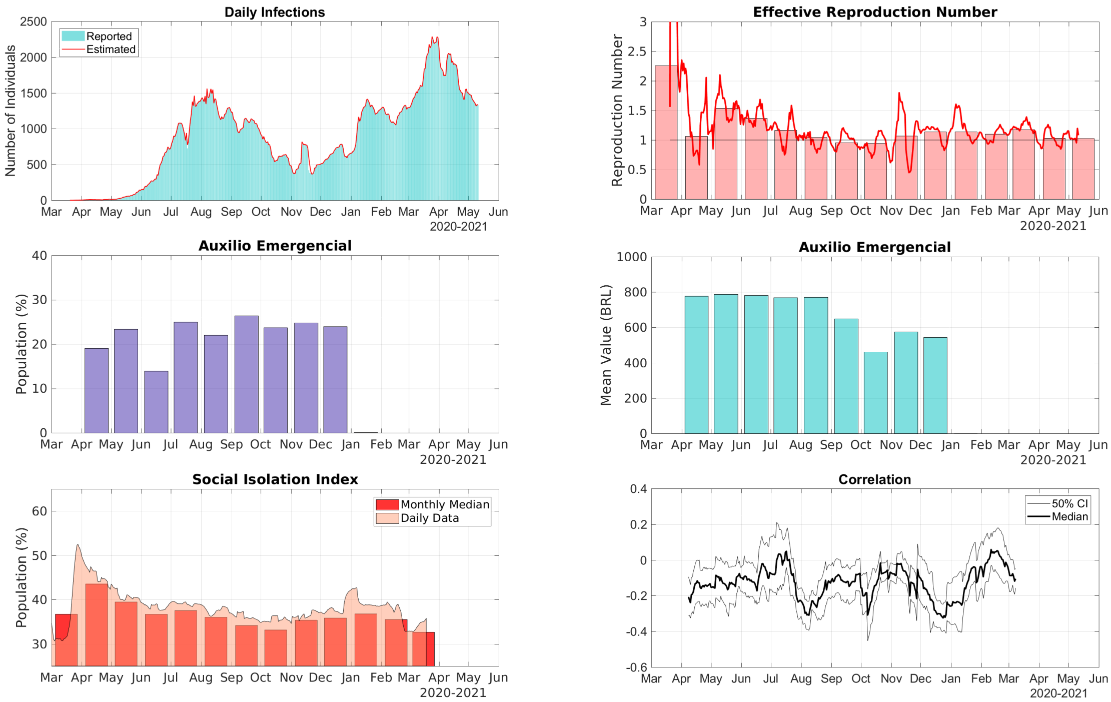


Figure S.19: Data for the State of Mato Grosso in the Central-West Region of Brazil. Top Row, Right: comparison between the 7-day moving average of daily reports of infections and model predictions (right). Top Row, Left: The solid line represents the time-dependent reproduction number and the bars are the corresponding monthly median values. Middle Row, Right: The bars represent the proportions of the State population receiving the Auxílio Emergencial by month. Middle Row, Left: The bars represent the statewide average amount paid by Auxílio Emergencial each month. Bottom Row, Left: The area graph is the 7-day moving average of the social isolation index and the bars represent the corresponding monthly median values. Bottom Row, Right: Correlation between the daily increments of the social isolation index and the reproduction number.

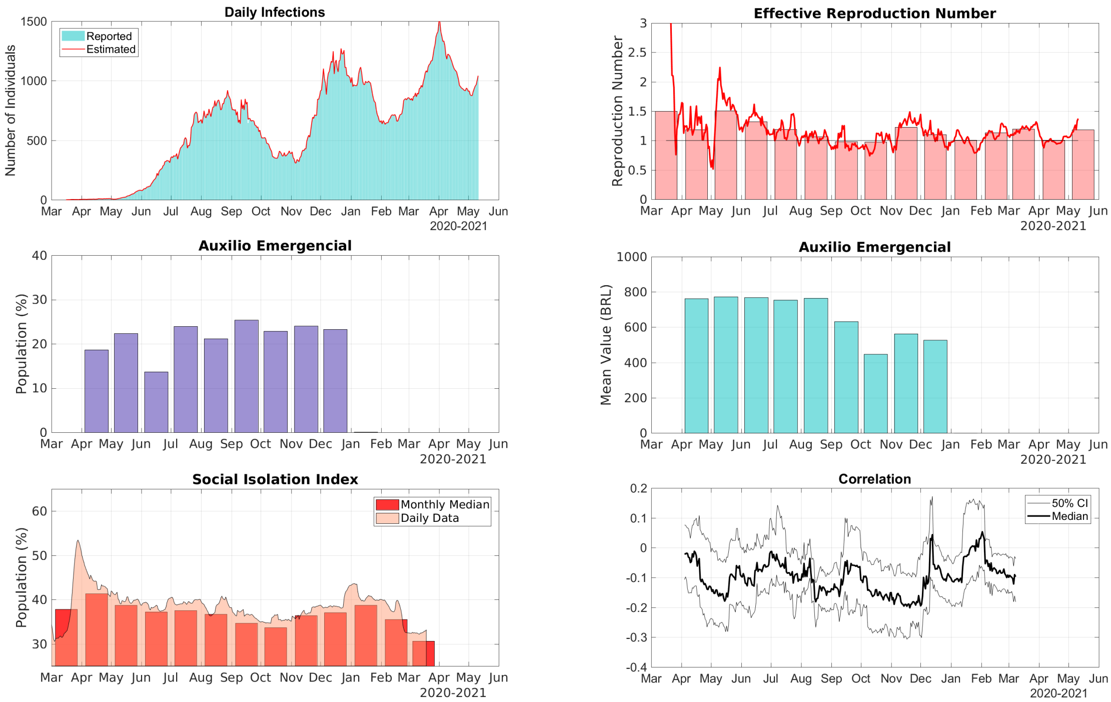


Figure S.20: Data for the State of Mato Grosso do Sul in the Central-West Region of Brazil. Top Row, Right: comparison between the 7-day moving average of daily reports of infections and model predictions (right). Top Row, Left: The solid line represents the time-dependent reproduction number and the bars are the corresponding monthly median values. Middle Row, Right: The bars represent the proportions of the State population receiving the Auxílio Emergencial by month. Middle Row, Left: The bars represent the statewide average amount paid by Auxílio Emergencial each month. Bottom Row, Left: The area graph is the 7-day moving average of the social isolation index and the bars represent the corresponding monthly median values. Bottom Row, Right: Correlation between the daily increments of the social isolation index and the reproduction number.

### 4 Southeast Region

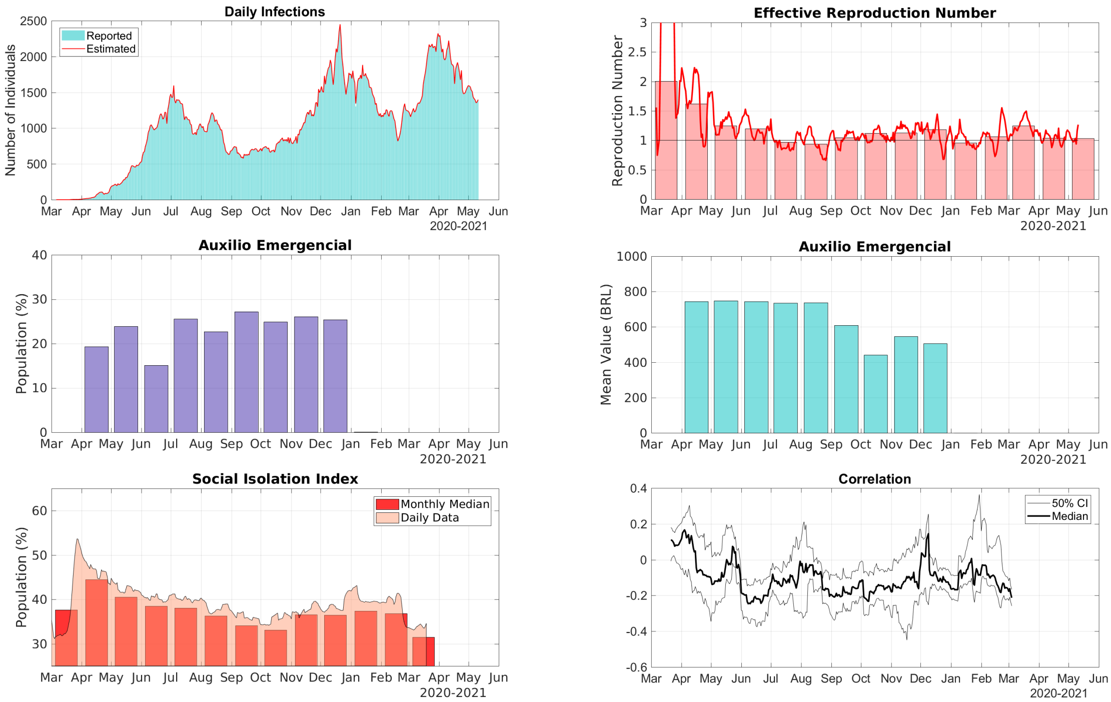


Figure S.21: Data for the State of Espírito Santo in the Southeast Region of Brazil. Top Row, Right: comparison between the 7-day moving average of daily reports of infections and model predictions (right). Top Row, Left: The solid line represents the time-dependent reproduction number and the bars are the corresponding monthly median values. Middle Row, Right: The bars represent the proportions of the State population receiving the Auxílio Emergencial by month. Middle Row, Left: The bars represent the statewide average amount paid by Auxílio Emergencial each month. Bottom Row, Left: The area graph is the 7-day moving average of the social isolation index and the bars represent the corresponding monthly median values. Bottom Row, Right: Correlation between the daily increments of the social isolation index and the reproduction number.

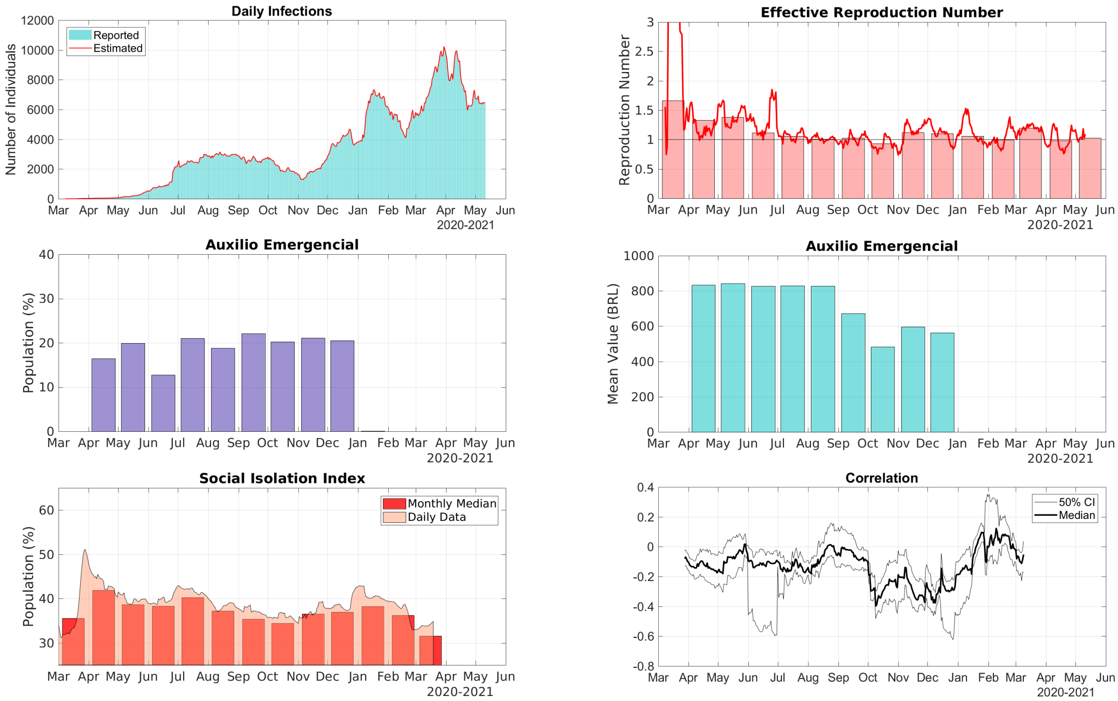


Figure S.22: Data for the State of Minas Gerais in the Southeast Region of Brazil. Top Row, Right: comparison between the 7-day moving average of daily reports of infections and model predictions (right). Top Row, Left: The solid line represents the time-dependent reproduction number and the bars are the corresponding monthly median values. Middle Row, Right: The bars represent the proportions of the State population receiving the Auxílio Emergencial by month. Middle Row, Left: The bars represent the statewide average amount paid by Auxílio Emergencial each month. Bottom Row, Left: The area graph is the 7-day moving average of the social isolation index and the bars represent the corresponding monthly median values. Bottom Row, Right: Correlation between the daily increments of the social isolation index and the reproduction number.

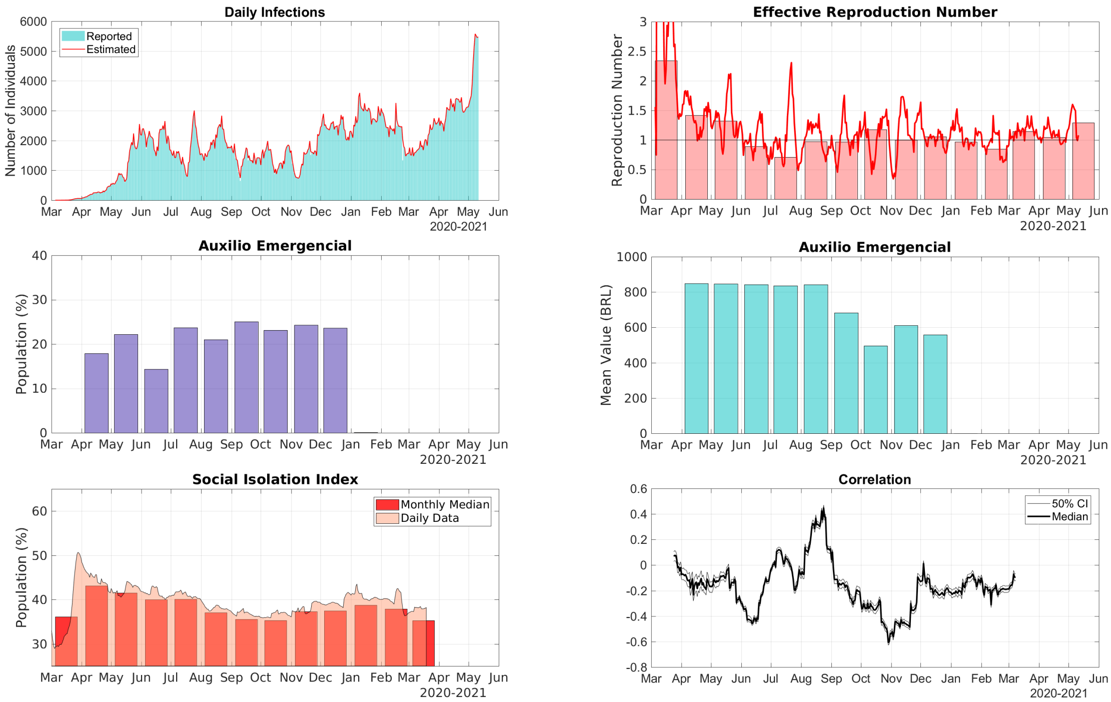


Figure S.23: Data for the State of Rio de Janeiro in the Southeast Region of Brazil. Top Row, Right: comparison between the 7-day moving average of daily reports of infections and model predictions (right). Top Row, Left: The solid line represents the time-dependent reproduction number and the bars are the corresponding monthly median values. Middle Row, Right: The bars represent the proportions of the State population receiving the Auxílio Emergencial by month. Middle Row, Left: The bars represent the statewide average amount paid by Auxílio Emergencial each month. Bottom Row, Left: The area graph is the 7-day moving average of the social isolation index and the bars represent the corresponding monthly median values. Bottom Row, Right: Correlation between the daily increments of the social isolation index and the reproduction number.

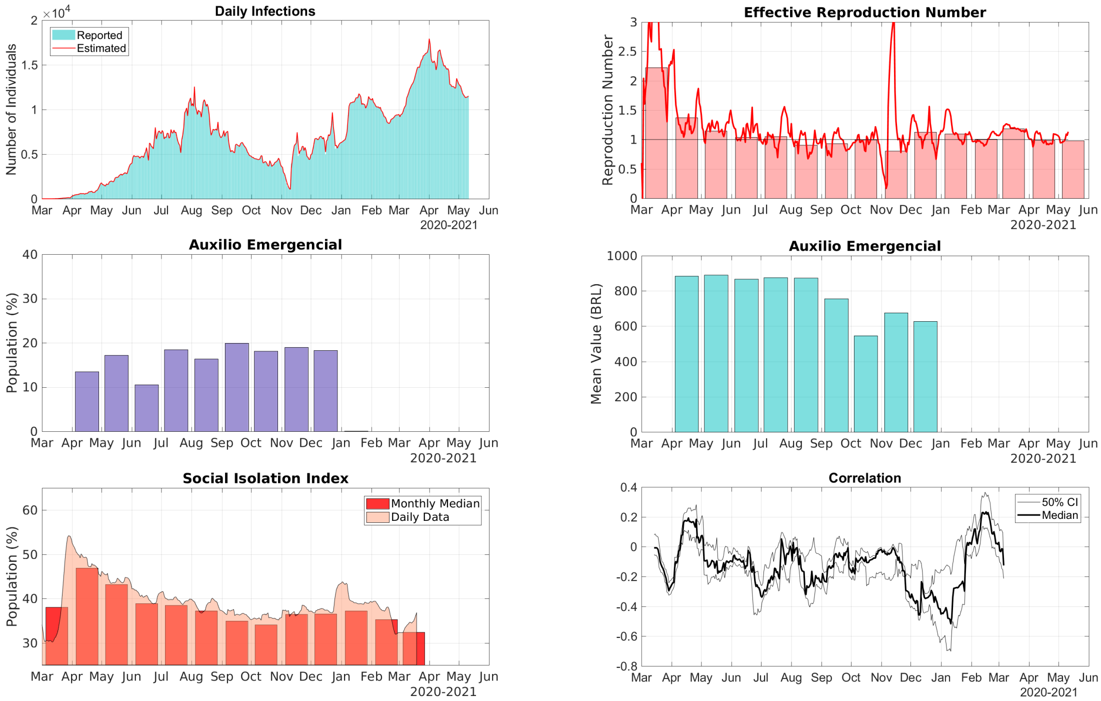


Figure S.24: Data for the State of São Paulo in the Southeast Region of Brazil. Top Row, Right: comparison between the 7-day moving average of daily reports of infections and model predictions (right). Top Row, Left: The solid line represents the time-dependent reproduction number and the bars are the corresponding monthly median values. Middle Row, Right: The bars represent the proportions of the State population receiving the Auxílio Emergencial by month. Middle Row, Left: The bars represent the statewide average amount paid by Auxílio Emergencial each month. Bottom Row, Left: The area graph is the 7-day moving average of the social isolation index and the bars represent the corresponding monthly median values. Bottom Row, Right: Correlation between the daily increments of the social isolation index and the reproduction number.

### 5 South Region

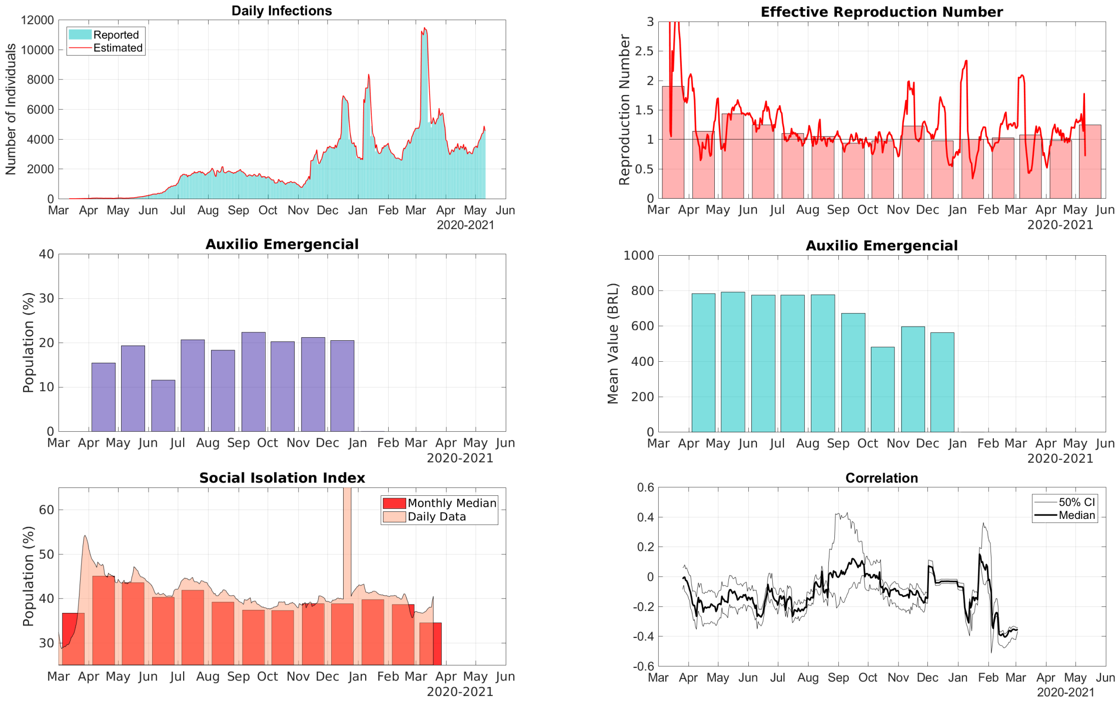


Figure S.25: Data for the State of Paraná in the South Region of Brazil. Top Row, Right: comparison between the 7-day moving average of daily reports of infections and model predictions (right). Top Row, Left: The solid line represents the time-dependent reproduction number and the bars are the corresponding monthly median values. Middle Row, Right: The bars represent the proportions of the State population receiving the Auxílio Emergencial by month. Middle Row, Left: The bars represent the statewide average amount paid by Auxílio Emergencial each month. Bottom Row, Left: The area graph is the 7-day moving average of the social isolation index and the bars represent the corresponding monthly median values. Bottom Row, Right: Correlation between the daily increments of the social isolation index and the reproduction number.

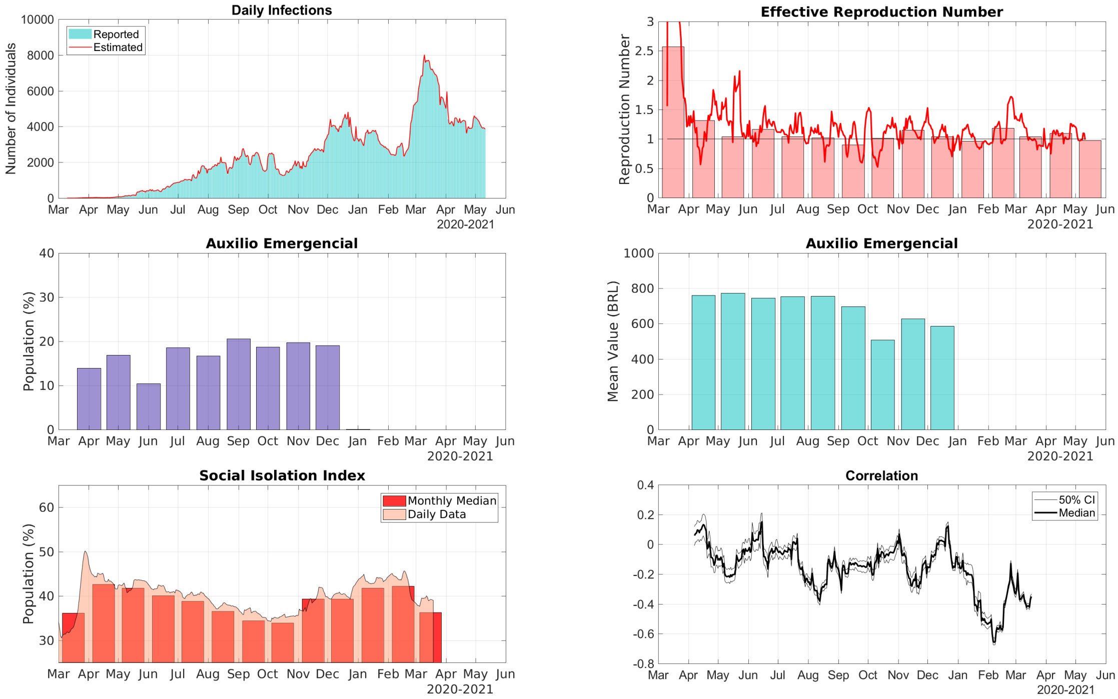


Figure S.26: Data for the State of Rio Grande do Sul in the South Region of Brazil. Top Row, Right: comparison between the 7-day moving average of daily reports of infections and model predictions (right). Top Row, Left: The solid line represents the time-dependent reproduction number and the bars are the corresponding monthly median values. Middle Row, Right: The bars represent the proportions of the State population receiving the Auxílio Emergencial by month. Middle Row, Left: The bars represent the statewide average amount paid by Auxílio Emergencial each month. Bottom Row, Left: The area graph is the 7-day moving average of the social isolation index and the bars represent the corresponding monthly median values. Bottom Row, Right: Correlation between the daily increments of the social isolation index and the reproduction number.

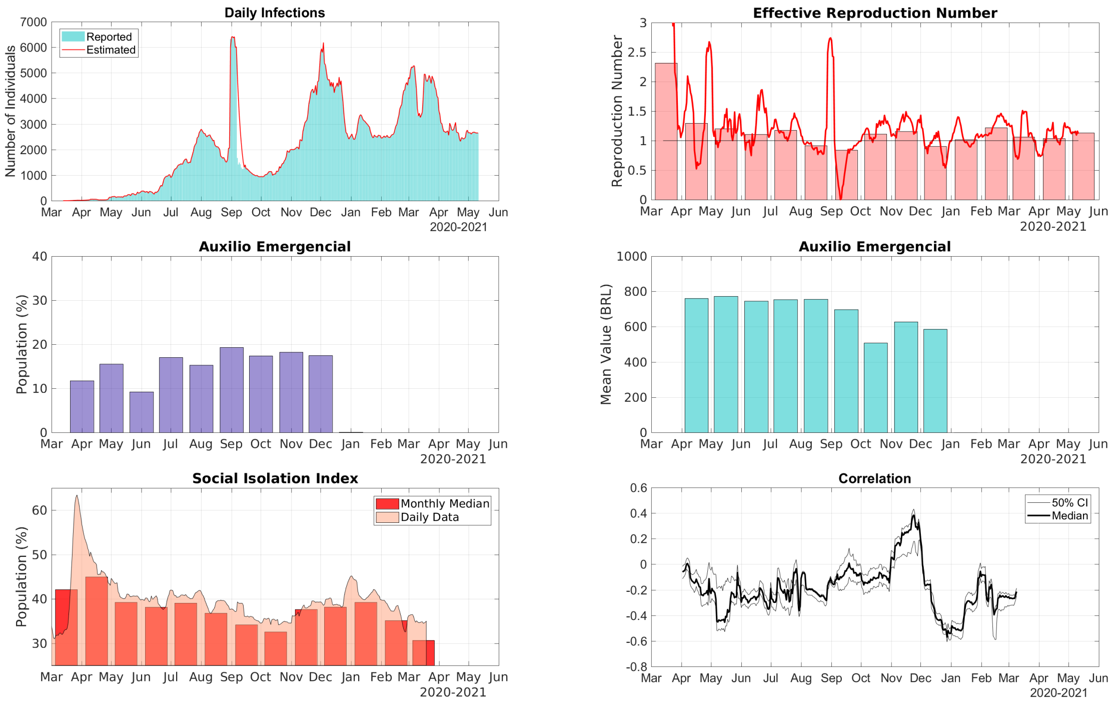


Figure S.27: Data for the State of Santa Catarina in the South Region of Brazil. Top Row, Right: comparison between the 7-day moving average of daily reports of infections and model predictions (right). Top Row, Left: The solid line represents the time-dependent reproduction number and the bars are the corresponding monthly median values. Middle Row, Right: The bars represent the proportions of the State population receiving the Auxílio Emergencial by month. Middle Row, Left: The bars represent the statewide average amount paid by Auxílio Emergencial each month. Bottom Row, Left: The area graph is the 7-day moving average of the social isolation index and the bars represent the corresponding monthly median values. Bottom Row, Right: Correlation between the daily increments of the social isolation index and the reproduction number.

### 6 Additional Tables

| State | Region | Mar-20 | Apr-20 | May-20 | Jun-20 | Jul-20 | Aug-20 | Sep-20 | Oct-20 | Nov-20 | Dec-20 | Jan-21 | Feb-21 | Mar-21 | Apr-21 | May-21 |
|---|---|---|---|---|---|---|---|---|---|---|---|---|---|---|---|---|
| AC | N | 3,06 | 1,52 | 1,36 | 1,02 | 0,99 | 0,98 | 1,03 | 1,00 | 1,19 | 1,06 | 1,22 | 1,15 | 1,10 | 0,84 | 0,89 |
| AL | NE | 1,10 | 2,09 | 1,42 | 1,06 | 0,96 | 0,81 | 0,91 | 0,90 | 1,15 | 1,16 | 1,11 | 0,95 | 1,17 | 1,08 | 1,12 |
| AM | N | 2,38 | 1,68 | 1,30 | 0,93 | 1,11 | 1,01 | 1,07 | 0,96 | 0,96 | 1,10 | 1,19 | 0,94 | 1,08 | 1,02 | 1,05 |
| AP | N | 1,39 | 1,82 | 1,44 | 1,00 | 0,93 | 1,13 | 0,95 | 1,07 | 1,26 | 1,07 | 1,11 | 1,08 | 1,34 | 0,90 | 1,20 |
| BA | NE | 1,58 | 1,30 | 1,28 | 1,23 | 1,00 | 1,00 | 0,92 | 1,04 | 1,18 | 1,09 | 1,04 | 1,12 | 1,01 | 1,05 | 1,09 |
| CE | NE | 1,78 | 1,54 | 1,26 | 0,99 | 1,02 | 0,89 | 0,88 | 0,97 | 1,07 | 0,93 | 1,13 | 1,02 | 1,12 | 0,95 | 1,05 |
| DF | CW | 1,93 | 1,06 | 1,41 | 1,37 | 1,05 | 1,00 | 0,95 | 1,04 | 1,06 | 1,16 | 1,16 | 1,18 | 1,16 | 1,04 | 1,12 |
| ES | SE | 2,00 | 1,62 | 1,25 | 1,19 | 0,97 | 0,94 | 1,05 | 1,12 | 1,13 | 1,19 | 0,96 | 1,07 | 1,25 | 1,05 | 1,03 |
| GO | CW | 1,48 | 1,51 | 1,24 | 1,29 | 1,05 | 1,05 | 0,98 | 0,84 | 0,97 | 0,98 | 1,13 | 1,10 | 1,15 | 1,06 | 0,89 |
| MA | NE | 3,00 | 1,62 | 1,52 | 0,92 | 1,09 | 1,01 | 0,92 | 0,91 | 1,04 | 1,01 | 1,11 | 1,13 | 1,13 | 1,03 | 1,04 |
| MG | SE | 1,66 | 1,33 | 1,38 | 1,11 | 1,06 | 1,01 | 1,03 | 0,93 | 1,12 | 1,11 | 1,06 | 0,99 | 1,20 | 0,99 | 1,03 |
| MS | CW | 1,50 | 1,19 | 1,51 | 1,32 | 1,19 | 1,07 | 0,98 | 0,98 | 1,23 | 1,11 | 1,01 | 1,14 | 1,20 | 1,01 | 1,19 |
| MT | CW | 2,26 | 1,06 | 1,54 | 1,37 | 1,17 | 1,05 | 0,96 | 0,95 | 1,07 | 1,14 | 1,15 | 1,11 | 1,19 | 1,04 | 1,04 |
| PA | N | 3,05 | 1,76 | 1,50 | 1,02 | 0,91 | 1,03 | 0,90 | 0,93 | 1,00 | 1,05 | 1,20 | 1,07 | 1,09 | 1,01 | 1,02 |
| PB | NE | 1,56 | 1,67 | 1,55 | 1,11 | 1,01 | 0,93 | 0,94 | 1,04 | 1,12 | 1,15 | 1,15 | 1,11 | 1,08 | 1,03 | 1,10 |
| PE | NE | 1,16 | 1,91 | 1,13 | 0,97 | 1,07 | 0,93 | 0,89 | 1,01 | 1,14 | 1,10 | 1,07 | 0,99 | 1,02 | 1,10 | 1,14 |
| PI | NE | 2,02 | 1,74 | 1,26 | 1,25 | 1,07 | 0,95 | 0,98 | 1,00 | 1,08 | 1,09 | 1,08 | 1,09 | 1,21 | 1,04 | 1,08 |
| PR | S | 1,90 | 1,14 | 1,44 | 1,25 | 1,10 | 1,05 | 0,94 | 0,98 | 1,23 | 0,98 | 1,00 | 1,03 | 1,08 | 0,98 | 1,25 |
| RJ | SE | 2,34 | 1,42 | 1,32 | 0,89 | 0,71 | 0,98 | 0,97 | 1,18 | 1,00 | 1,06 | 0,97 | 0,85 | 1,15 | 1,05 | 1,30 |
| RN | NE | 1,86 | 1,27 | 1,22 | 1,31 | 0,72 | 0,93 | 0,95 | 1,00 | 1,27 | 1,14 | 1,04 | 0,96 | 1,12 | 1,24 | 1,05 |
| RO | N | 1,40 | 1,96 | 1,41 | 1,31 | 0,78 | 0,99 | 0,91 | 0,90 | 1,26 | 1,15 | 1,21 | 1,07 | 1,16 | 1,02 | 1,14 |
| RR | N | 2,25 | 1,55 | 1,19 | 1,37 | 0,91 | 1,03 | 1,09 | 1,14 | 0,91 | 0,88 | 1,33 | 1,23 | 1,10 | 1,26 | 1,28 |
| RS | S | 2,57 | 1,31 | 1,04 | 1,17 | 1,04 | 1,02 | 0,90 | 1,01 | 1,15 | 1,04 | 0,96 | 1,18 | 1,04 | 1,10 | 0,98 |
| SC | S | 2,32 | 1,29 | 1,20 | 1,11 | 1,18 | 0,92 | 0,84 | 1,11 | 1,16 | 0,91 | 1,02 | 1,23 | 1,07 | 1,05 | 1,15 |
| SE | NE | 1,03 | 1,68 | 1,44 | 1,22 | 1,12 | 0,83 | 0,96 | 1,15 | 1,41 | 1,27 | 1,09 | 1,01 | 1,30 | 1,03 | 1,06 |
| SP | SE | 2,23 | 1,38 | 1,15 | 1,04 | 1,05 | 0,91 | 0,93 | 0,99 | 0,81 | 1,13 | 1,10 | 1,01 | 1,18 | 0,99 | 0,98 |
| TO | N | 1,24 | 1,36 | 1,54 | 1,03 | 1,11 | 1,11 | 0,92 | 0,86 | 1,16 | 1,07 | 1,08 | 1,12 | 1,18 | 1,05 | 1,12 |

Table S.1: Monthly median values of the reproduction number in the Brazilian states.

| State | Region | Apr-20 | May-20 | Jun-20 | Jul-20 | Aug-20 | Sep-20 | Oct-20 | Nov-20 | Dec-20 |
|---|---|---|---|---|---|---|---|---|---|---|
| AC | N | R$ 752,28 | R$ 755,60 | R$ 757,13 | R$ 747,78 | R$ 750,59 | R$ 501,52 | R$ 365,14 | R$ 444,98 | R$ 416,38 |
| AL | NE | R$ 985,97 | R$ 982,85 | R$ 993,75 | R$ 970,99 | R$ 974,97 | R$ 627,79 | R$ 461,30 | R$ 556,13 | R$ 528,66 |
| AM | N | R$ 780,90 | R$ 780,21 | R$ 786,16 | R$ 772,84 | R$ 778,24 | R$ 545,09 | R$ 400,33 | R$ 483,18 | R$ 451,91 |
| AP | N | R$ 778,26 | R$ 775,91 | R$ 795,34 | R$ 766,05 | R$ 770,25 | R$ 558,66 | R$ 397,73 | R$ 491,10 | R$ 463,51 |
| BA | NE | R$ 937,55 | R$ 941,72 | R$ 940,22 | R$ 931,73 | R$ 932,72 | R$ 612,42 | R$ 445,48 | R$ 542,90 | R$ 519,31 |
| CE | NE | R$ 846,61 | R$ 848,23 | R$ 851,73 | R$ 839,80 | R$ 841,70 | R$ 548,83 | R$ 399,18 | R$ 484,63 | R$ 464,09 |
| DF | CW | R$ 769,64 | R$ 769,37 | R$ 762,81 | R$ 754,89 | R$ 761,52 | R$ 662,92 | R$ 482,11 | R$ 593,56 | R$ 557,28 |
| ES | SE | R$ 744,43 | R$ 748,46 | R$ 744,19 | R$ 735,21 | R$ 736,41 | R$ 608,66 | R$ 441,69 | R$ 546,38 | R$ 506,08 |
| GO | CW | R$ 801,76 | R$ 807,14 | R$ 797,68 | R$ 791,64 | R$ 792,03 | R$ 671,92 | R$ 483,87 | R$ 594,82 | R$ 562,63 |
| MA | NE | R$ 866,46 | R$ 871,20 | R$ 877,13 | R$ 858,27 | R$ 861,45 | R$ 531,70 | R$ 390,30 | R$ 467,19 | R$ 458,47 |
| MG | SE | R$ 834,10 | R$ 842,36 | R$ 827,68 | R$ 829,24 | R$ 828,48 | R$ 671,52 | R$ 482,11 | R$ 597,12 | R$ 562,48 |
| MS | CW | R$ 762,56 | R$ 773,00 | R$ 768,47 | R$ 755,02 | R$ 765,51 | R$ 632,85 | R$ 446,42 | R$ 562,13 | R$ 526,91 |
| MT | CW | R$ 778,40 | R$ 788,24 | R$ 782,33 | R$ 769,53 | R$ 771,59 | R$ 649,78 | R$ 461,15 | R$ 575,00 | R$ 544,76 |
| PA | N | R$ 809,95 | R$ 813,07 | R$ 819,26 | R$ 805,00 | R$ 807,85 | R$ 547,54 | R$ 396,53 | R$ 482,84 | R$ 463,57 |
| PB | NE | R$ 859,16 | R$ 859,63 | R$ 865,49 | R$ 850,28 | R$ 853,08 | R$ 533,89 | R$ 394,13 | R$ 475,56 | R$ 450,14 |
| PE | NE | R$ 990,51 | R$ 988,95 | R$ 999,31 | R$ 976,53 | R$ 983,44 | R$ 664,51 | R$ 474,76 | R$ 577,65 | R$ 548,07 |
| PI | NE | R$ 851,90 | R$ 855,71 | R$ 856,36 | R$ 846,75 | R$ 848,35 | R$ 522,08 | R$ 382,96 | R$ 461,44 | R$ 446,42 |
| PR | S | R$ 783,62 | R$ 792,75 | R$ 774,35 | R$ 775,62 | R$ 777,08 | R$ 672,41 | R$ 480,79 | R$ 597,13 | R$ 562,76 |
| RJ | SE | R$ 848,42 | R$ 846,16 | R$ 843,03 | R$ 835,77 | R$ 841,75 | R$ 682,27 | R$ 496,24 | R$ 610,48 | R$ 558,86 |
| RN | NE | R$ 832,20 | R$ 833,94 | R$ 837,32 | R$ 824,10 | R$ 829,27 | R$ 564,98 | R$ 406,54 | R$ 498,13 | R$ 472,49 |
| RO | N | R$ 743,91 | R$ 755,35 | R$ 746,47 | R$ 738,25 | R$ 740,95 | R$ 620,22 | R$ 438,58 | R$ 547,53 | R$ 515,04 |
| RR | N | R$ 748,40 | R$ 753,87 | R$ 750,43 | R$ 739,63 | R$ 746,65 | R$ 564,87 | R$ 412,72 | R$ 510,46 | R$ 469,91 |
| RS | S | R$ 771,56 | R$ 777,97 | R$ 769,12 | R$ 763,45 | R$ 774,51 | R$ 681,96 | R$ 475,20 | R$ 593,87 | R$ 557,20 |
| SC | S | R$ 761,53 | R$ 773,25 | R$ 745,64 | R$ 754,56 | R$ 756,38 | R$ 698,06 | R$ 507,44 | R$ 627,98 | R$ 586,93 |
| SE | NE | R$ 914,49 | R$ 913,83 | R$ 924,91 | R$ 904,31 | R$ 908,71 | R$ 594,17 | R$ 433,47 | R$ 527,89 | R$ 504,25 |
| SP | SE | R$ 885,30 | R$ 891,15 | R$ 868,12 | R$ 876,77 | R$ 873,79 | R$ 755,58 | R$ 546,93 | R$ 675,85 | R$ 627,22 |
| TO | N | R$ 753,58 | R$ 761,15 | R$ 767,12 | R$ 748,91 | R$ 751,80 | R$ 562,60 | R$ 392,69 | R$ 484,97 | R$ 475,22 |

Table S.2: Monthly average amount paid by Auxílio Emergencial during 2020 for each Brazilian state.

| State | Region | Feb-20 | Mar-20 | Apr-20 | May-20 | Jun-20 | Jul-20 | Aug-20 | Sep-20 | Oct-20 | Nov-20 | Dec-20 | Jan-21 | Feb-21 | Mar-21 |
|---|---|---|---|---|---|---|---|---|---|---|---|---|---|---|---|
| AC | N | 31,2% | 43,5% | 47,0% | 48,1% | 44,0% | 43,0% | 41,1% | 38,6% | 37,8% | 42,8% | 42,7% | 45,8% | 45,3% | 38,7% |
| AL | NE | 25,8% | 35,4% | 43,7% | 44,0% | 40,6% | 40,2% | 37,6% | 35,9% | 35,3% | 36,7% | 36,5% | 38,0% | 37,2% | 34,0% |
| AM | N | 27,8% | 42,6% | 47,7% | 51,8% | 43,7% | 41,6% | 38,7% | 37,3% | 36,2% | 39,9% | 39,8% | 42,9% | 42,0% | 36,3% |
| AP | N | 30,5% | 43,2% | 52,3% | 48,1% | 40,8% | 40,6% | 39,3% | 37,2% | 36,7% | 41,7% | 42,4% | 53,0% | 48,2% | 40,1% |
| BA | NE | 26,5% | 37,6% | 43,9% | 42,9% | 39,8% | 41,0% | 38,5% | 36,6% | 35,8% | 36,9% | 36,9% | 38,2% | 38,0% | 37,1% |
| CE | NE | 28,3% | 41,9% | 48,3% | 48,7% | 41,2% | 41,3% | 39,4% | 37,8% | 37,6% | 40,4% | 40,2% | 41,6% | 40,8% | 41,9% |
| DF | CW | 26,5% | 39,8% | 47,1% | 42,4% | 39,8% | 40,2% | 38,7% | 36,3% | 35,1% | 38,5% | 38,2% | 39,1% | 37,9% | 36,3% |
| ES | SE | 27,4% | 37,6% | 44,5% | 40,6% | 38,5% | 38,1% | 36,3% | 34,1% | 33,1% | 36,6% | 36,5% | 37,4% | 36,8% | 31,5% |
| GO | CW | 26,3% | 37,8% | 42,0% | 37,4% | 35,7% | 37,1% | 35,1% | 33,6% | 32,1% | 34,8% | 35,2% | 35,6% | 34,3% | 33,0% |
| MA | NE | 27,8% | 36,9% | 44,3% | 47,6% | 38,2% | 38,7% | 37,2% | 36,1% | 35,6% | 38,5% | 38,4% | 40,3% | 39,5% | 36,6% |
| MG | SE | 28,2% | 35,6% | 41,9% | 38,6% | 38,3% | 40,2% | 37,2% | 35,4% | 34,4% | 36,6% | 36,9% | 38,3% | 36,3% | 31,6% |
| MS | CW | 28,5% | 37,8% | 41,4% | 38,7% | 37,3% | 37,6% | 36,7% | 34,7% | 33,7% | 36,4% | 37,1% | 38,8% | 35,5% | 30,6% |
| MT | CW | 27,0% | 36,8% | 43,6% | 39,5% | 36,7% | 37,6% | 36,1% | 34,2% | 33,2% | 35,4% | 35,9% | 36,8% | 35,6% | 32,7% |
| PA | N | 27,4% | 37,4% | 44,3% | 39,4% | 37,9% | 38,6% | 37,1% | 34,1% | 32,8% | 36,8% | 36,8% | 38,3% | 35,5% | 34,3% |
| PB | NE | 26,5% | 36,9% | 44,5% | 44,0% | 40,5% | 40,0% | 37,5% | 36,0% | 35,8% | 38,5% | 38,4% | 39,5% | 38,9% | 35,6% |
| PE | NE | 28,1% | 37,9% | 46,2% | 48,6% | 38,8% | 38,2% | 37,4% | 35,6% | 34,9% | 38,9% | 39,0% | 41,5% | 41,4% | 36,8% |
| PI | NE | 26,5% | 39,6% | 48,2% | 47,3% | 40,9% | 40,0% | 38,1% | 36,8% | 36,0% | 37,7% | 37,7% | 38,6% | 38,0% | 33,5% |
| PR | S | 26,4% | 36,7% | 45,1% | 43,6% | 40,3% | 41,9% | 39,2% | 37,4% | 37,3% | 38,9% | 38,8% | 39,8% | 38,7% | 34,5% |
| RJ | SE | 26,2% | 36,1% | 43,1% | 41,5% | 40,0% | 40,1% | 37,1% | 35,6% | 35,3% | 37,3% | 37,4% | 38,8% | 37,9% | 35,3% |
| RN | NE | 28,1% | 39,8% | 47,3% | 41,6% | 38,8% | 40,3% | 37,7% | 35,4% | 34,3% | 35,9% | 36,5% | 36,6% | 35,2% | 34,1% |
| RO | N | 29,2% | 41,1% | 49,5% | 46,6% | 41,3% | 40,3% | 38,6% | 37,1% | 35,8% | 39,3% | 39,9% | 40,6% | 39,2% | 33,2% |
| RR | N | 29,4% | 38,7% | 44,6% | 43,0% | 42,0% | 41,3% | 38,9% | 36,9% | 36,4% | 40,3% | 40,5% | 46,2% | 42,3% | 37,6% |
| RS | S | 27,3% | 36,2% | 42,7% | 41,8% | 40,1% | 38,8% | 36,6% | 34,4% | 33,9% | 39,4% | 39,3% | 41,8% | 42,2% | 36,3% |
| SC | S | 28,5% | 42,1% | 45,0% | 39,3% | 38,1% | 39,1% | 36,8% | 34,2% | 32,6% | 37,6% | 38,2% | 39,3% | 35,2% | 30,7% |
| SE | NE | 26,0% | 36,7% | 42,3% | 41,6% | 38,3% | 40,1% | 37,5% | 35,2% | 34,2% | 37,3% | 36,7% | 38,1% | 37,2% | 35,0% |
| SP | SE | 26,5% | 38,1% | 46,9% | 43,2% | 38,9% | 38,5% | 37,3% | 35,0% | 34,1% | 36,5% | 36,6% | 37,3% | 35,3% | 32,4% |
| TO | N | 24,6% | 38,3% | 38,4% | 37,5% | 34,4% | 35,1% | 34,7% | 33,2% | 31,3% | 33,4% | 33,8% | 34,9% | 33,5% | 32,5% |

Table S.3: Monthly median values of the social isolation index (SII).

| State | Region | Feb-20 | Mar-20 | Apr-20 | May-20 | Jun-20 | Jul-20 | Aug-20 | Sep-20 | Oct-20 | Nov-20 | Dec-20 | Jan-21 | Feb-21 | Mar-21 | Apr-21 |
|---|---|---|---|---|---|---|---|---|---|---|---|---|---|---|---|---|
| AC | N | 0,00 | 4,70 | 40,5 | 650 | 786 | 712 | 561 | 400 | 288 | 611 | 599 | 765 | 1014 | 1355 | 911 |
| AL | NE | 0,00 | 0,54 | 30,6 | 276 | 766 | 709 | 569 | 241 | 117 | 125 | 293 | 386 | 417 | 649 | 602 |
| AM | N | 0,00 | 4,16 | 121 | 859 | 700 | 716 | 460 | 452 | 531 | 393 | 542 | 1578 | 1154 | 788 | 515 |
| AP | N | 0,00 | 1,16 | 124 | 989 | 2192 | 926 | 780 | 587 | 455 | 807 | 1052 | 1026 | 768 | 1611 | 953 |
| BA | NE | 0,00 | 1,43 | 17,7 | 104 | 368 | 622 | 607 | 360 | 286 | 334 | 605 | 634 | 643 | 801 | 651 |
| CE | NE | 0,00 | 4,25 | 78,5 | 445 | 655 | 710 | 447 | 285 | 359 | 288 | 375 | 417 | 566 | 1229 | 1407 |
| DF | CW | 0,00 | 10,9 | 33,5 | 276 | 1291 | 1868 | 1825 | 989 | 687 | 520 | 738 | 832 | 641 | 1560 | 1127 |
| ES | SE | 0,00 | 2,07 | 58,6 | 276 | 817 | 896 | 688 | 498 | 596 | 841 | 1436 | 1128 | 792 | 1375 | 1339 |
| GO | CW | 0,00 | 0,91 | 10,1 | 41,1 | 291 | 612 | 908 | 1081 | 647 | 337 | 414 | 586 | 636 | 1244 | 944 |
| MA | NE | 0,00 | 0,44 | 44,4 | 442 | 644 | 565 | 435 | 309 | 172 | 103 | 109 | 92 | 166 | 325 | 344 |
| MG | SE | 0,00 | 1,29 | 7,29 | 40,6 | 162 | 386 | 420 | 369 | 300 | 269 | 594 | 900 | 677 | 1152 | 1105 |
| MS | CW | 0,00 | 1,71 | 7,37 | 43,9 | 231 | 604 | 854 | 739 | 450 | 595 | 1235 | 970 | 730 | 1213 | 1160 |
| MT | CW | 0,00 | 0,71 | 7,71 | 60,5 | 375 | 1027 | 1120 | 909 | 553 | 432 | 604 | 1032 | 953 | 1677 | 1457 |
| PA | N | 0,00 | 0,37 | 32,7 | 404 | 751 | 592 | 516 | 355 | 259 | 203 | 265 | 412 | 407 | 608 | 622 |
| PB | NE | 0,00 | 0,42 | 19,7 | 306 | 837 | 887 | 569 | 383 | 294 | 300 | 526 | 621 | 731 | 942 | 829 |
| PE | NE | 0,00 | 0,90 | 70,6 | 287 | 254 | 376 | 318 | 225 | 163 | 203 | 413 | 407 | 397 | 517 | 596 |
| PI | NE | 0,00 | 0,55 | 15,1 | 135 | 472 | 946 | 790 | 569 | 524 | 418 | 477 | 498 | 447 | 954 | 1079 |
| PR | S | 0,00 | 1,55 | 10,7 | 28,5 | 158 | 462 | 484 | 408 | 300 | 578 | 1185 | 1153 | 848 | 1729 | 882 |
| RJ | SE | 0,00 | 4,08 | 50,4 | 253 | 341 | 305 | 335 | 237 | 260 | 256 | 462 | 497 | 357 | 373 | 546 |
| RN | NE | 0,00 | 2,32 | 31,0 | 176 | 648 | 570 | 321 | 217 | 332 | 395 | 650 | 637 | 745 | 831 | 756 |
| RO | N | 0,00 | 0,45 | 27,5 | 247 | 908 | 988 | 900 | 599 | 320 | 494 | 846 | 1611 | 1342 | 2143 | 1418 |
| RR | N | 0,00 | 2,53 | 79,7 | 468 | 1769 | 2753 | 1822 | 1098 | 1083 | 957 | 850 | 860 | 1257 | 1184 | 1076 |
| RS | S | 0,00 | 2,40 | 10,4 | 68,9 | 154 | 348 | 518 | 567 | 499 | 652 | 1117 | 854 | 820 | 1790 | 1134 |
| SC | S | 0,00 | 3,02 | 25,7 | 95,9 | 239 | 796 | 1292 | 520 | 599 | 1453 | 1768 | 1161 | 1293 | 1880 | 1117 |
| SE | NE | 0,00 | 0,82 | 18,5 | 283 | 794 | 1436 | 596 | 212 | 297 | 251 | 964 | 1074 | 603 | 1000 | 1172 |
| SP | SE | 0,00 | 5,05 | 56,9 | 175 | 371 | 564 | 566 | 392 | 282 | 271 | 477 | 681 | 571 | 925 | 937 |
| TO | N | 0,00 | 0,69 | 7,92 | 254 | 414 | 884 | 1627 | 1088 | 476 | 381 | 549 | 746 | 736 | 1701 | 1178 |

Table S.4: Number of COVID-19 infections per 100K individuals by month for each Brazilian state.
